# Screening for SARS-CoV-2 in close contacts of individuals with confirmed infection: performance and operational considerations

**DOI:** 10.1101/2022.01.27.22269904

**Authors:** Stephanie Zobrist, Michelle Oliveira-Silva, Alexia Martines Vieira, Pooja Bansil, Emily Gerth-Guyette, Brandon T. Leader, Allison Golden, Hannah Slater, Catherine Duran de Lucena Cruz, Eduardo Garbin, Mariana Sagalovsky, Sampa Pal, Vin Gupta, Leo Wolansky, Deusilene Souza Vieira Dall’Acqua, Felipe Gomes Naveca, Valdinete Alves do Nascimento, Juan Miguel Villalobos Salcedo, Paul K. Drain, Alexandre Dias Tavares Costa, Gonzalo J Domingo, Dhélio Pereira

## Abstract

**Background:** Point-of-care and decentralized testing for SARS-CoV-2 is critical to inform public health responses. Performance evaluations in priority use cases such as contact tracing can highlight trade-offs in test selection and testing strategies.

**Methods:** A prospective diagnostic accuracy study was conducted among close contacts of COVID-19 cases in Brazil. Two anterior nares swabs (ANS), a nasopharyngeal swab (NPS), and saliva were collected at all visits. Vaccination history and symptoms were assessed. Household contacts were followed longitudinally. Three rapid antigen tests and one molecular method were evaluated for usability and performance against reference RT-PCR on NPS.

**Results:** Fifty index cases and 214 contacts (64 household) were enrolled. Sixty-five contacts were RT-PCR positive during at least one visit. Vaccination did not influence viral load. Gamma variants were most prevalent; Delta emerged increasingly during implementation. Overall sensitivity of evaluated tests ranged from 33%–76%. Performance was higher among symptomatic cases and cases with Ct<34 and lower among oligo/asymptomatic cases. Assuming a 24-hour time-to-result for RT-PCR, the cumulative sensitivity of an ANS rapid antigen test was >70% and almost 90% after four days.

**Conclusions:** The near immediate time-to-result for antigen tests significantly offsets lower analytical sensitivity in settings where RT-PCR results are delayed or unavailable.

## Introduction

The severe acute respiratory syndrome coronavirus 2 (SARS-CoV-2) virus, which causes COVID-19, has significantly burdened health systems globally, with over 22 million confirmed cases in Brazil alone as of 2021 [1]. A key challenge of the pandemic response is access to appropriate diagnostic testing, which is critical to inform public health strategies [2].

The reference standard for SARS-CoV-2 testing is RT-PCR. While accurate, this method has many practical limitations, including cost, laboratory infrastructure requirements, and often invasive sampling. RT-PCR testing is typically centralized, which can lead to delays in reporting results to patients. Such delays have important public health implications, including increased risk for transmission during the period before results are available [3,4]. Expanded access to decentralized and point-of-care (POC) testing is essential to identify cases early and limit community transmission, particularly where RT-PCR is unavailable.

Infected persons both with and without symptoms can transmit SARS-CoV-2 [5–7]. Due to the significance of asymptomatic transmission [8], testing these populations is often recommended, including close contacts of individuals with confirmed infection as part of contact tracing, testing, and isolation strategies [9,10].

Multiple platforms have been developed to enable decentralized and POC SARS-CoV-2 testing [11]. In particular, rapid antigen tests have garnered interest due to their lower cost, ease of use, and rapid turnaround time for results (typically <30 minutes) [10, 12]. The World Health Organization (WHO) advises that rapid antigen tests meeting minimum performance criteria can be employed in a range of use cases, including for testing of asymptomatic contacts of cases [10]. Previous performance evaluations have shown variability, with strongest performance among symptomatic individuals with high viral loads in early stages of infection [11, 13–15]. Several studies have investigated test performance among contacts of confirmed cases [16,17]; however, more data are needed to understand trade-offs in test selection and inform screening strategies regarding the timing and frequency of testing and performance characteristics.

## Methods

### Study design and population

A prospective diagnostic accuracy study was conducted among close contacts of COVID-19-positive index cases in Porto Velho, Brazil, between July and September 2021. Symptomatic adults within seven days of symptom onset who tested positive on a rapid SARS-CoV-2 antigen test (STANDARD Q COVID-19 Ag Nasal Test, SD Biosensor, Republic of Korea) were recruited as index cases. Close contacts were identified through interviews administered at enrollment of the index case. Individuals 12 years of age or older who resided in Porto Velho were eligible for inclusion as close contacts if they met one or more of Brazil’s criteria within the investigation period of the index case (two days prior to symptom onset to the time of the interview) (Supplementary Material A) [18]. Contacts with prior positive COVID-19 test results within the past three months were not eligible. A subset of household contacts (who shared a primary residence with the index case) had serial visits for clinical evaluations and testing every other day over nine days.

### Tests evaluated

This study evaluated four SARS-CoV-2 tests: the STANDARD Q COVID-19 Ag Nasal and Saliva tests, the SARS-CoV-2 Ag Test (LumiraDx™ Limited, United Kingdom), and the SalivaDirect™ protocol (Yale School of Public Health, United States). The STANDARD Q tests are rapid chromatographic immunoassays for qualitative detection of antigens from SARS-CoV-2 in human nasal and saliva specimens, respectively. The LumiraDx test is a microfluidic immunofluorescence assay for qualitative detection of antigen in nasal specimens [19–24]. SalivaDirect is a dual-plexed RT-PCR method for SARS-CoV-2 detection from minimally processed saliva [25,26].

### Study procedures at the point of care

At enrollment, information on participant demographics, health status, and medical history were collected. Presence, duration, and severity of symptoms were assessed at all visits, and two paired anterior nares swabs (ANS), one nasopharyngeal swab (NPS), and saliva were collected (Supplementary Material B). One ANS was used to run the STANDARD Q COVID-19 Ag Nasal Test during the visit. All specimens were then transferred to a laboratory where the remaining tests were performed.

For household contacts in the longitudinal sample, if the POC screening test was positive, only one additional study visit occurred, during which NPS were not collected to minimize staff exposure. Participants were considered lost to follow-up after two missed visits.

### Laboratory procedures

The extracted ANS mixed with LumiraDx buffer was frozen within five hours of collection and thawed before testing, no more than five days after freezing. The saliva sample was also frozen; aliquots were thawed for testing with the STANDARD Q COVID-19 Ag Saliva Test (within five days of freezing) and SalivaDirect. Evaluated tests were conducted per manufacturer instructions and by operators blinded to close contact POC and reference results.

#### Reference testing

NPS were used for reference testing with the Allplex™ SARS-CoV-2 Assay (Seegene Inc., Republic of Korea), a multiplex real-time PCR assay, on a CFX96 real-time PCR machine (Bio-Rad, United States) [27]. Automated RNA extraction was conducted using the Loccus Extracta kit (Loccus, Brazil). All SARS-CoV-2-positive specimens were repeated on the same assay for quantitative estimation of viral load. Specimens with cycle threshold (Ct) values <30 underwent genomic sequencing (Supplementary Material C). Staff conducting reference testing were blinded to other close contact test results.

### Usability assessment

Study staff responsible for use of the antigen tests were invited to participate in a usability assessment. A System Usability Scale (SUS) was employed, and an Ease of Use (EoU) questionnaire was adapted [21,28] (Supplementary Material D). SUS scores above 68 were considered acceptable [29,30]. To analyze data from the EoU questionnaire, a matrix was used to rank aspects of the products’ usability as “satisfactory,” “average,” or “unsatisfactory” (Supplementary Material E) [21].

### Sample size and statistical analysis

The sample size targeted at least 50 contacts with a positive reference result, including at least 20 asymptomatic individuals, to meet US FDA Emergency Use Authorization requirements [31].

Participants with no symptoms at the time of sampling were classified as asymptomatic. Participants were considered symptomatic if they presented with cough, shortness of breath, difficulty breathing, or at least two of the following symptoms at the time of sampling: fever, chills, rigor, myalgia, headache, sore throat, new olfactory or taste disorder [32]. Participants who presented with one or more mild symptoms but did not fit the symptomatic case definition and reported no care seeking or changes to behavior were considered oligosymptomatic.

Sensitivity, specificity, and positive and negative predictive values were calculated using standard formulas and presented with 95% CIs. Samples for which both RT-PCR and evaluated test results were available were included in the analysis. Using the longitudinal dataset, trade-offs between performance and utility of the evaluated tests in terms of cumulative sensitivity at specified time points were assessed as a function of time-to-results.

Data were collected and managed using REDCap electronic data capture tools hosted at the Institute of Translational Health Sciences [33]. Statistical analyses were conducted using Stata 15.0 (StataCorp, College Station, Texas, USA) and R 4.0.3 (R Foundation for Statistical Computing, Vienna, Austria).

### Ethical considerations

WCG Institutional Review Board (1301165), the CEPEM ethics committee, and Brazil’s National Research Ethics Commission approved this study (44351421.0.0000.0011). Written informed consent was obtained for all participants. Minors under 18 provided assent, and written informed consent was obtained from parents/legal guardians.

## Results

### Participant characteristics

Fifty symptomatic COVID-19-positive index cases and 214 of their associated close contacts were enrolled (Table 1). Sixty-four contacts shared a primary residence with an index case and were therefore included in the longitudinal sample. Contacts ranged from ages 13 to 79. The majority of participants across all groups were female. Sixty-five contacts (30%, 65/214) were SARS-CoV-2 positive by the reference assay during at least one visit (Figure 1). For household contacts, positivity rates and symptom status varied by visit. In total, 42 paired samples were collected at unique visits with oligo/asymptomatic positive contacts, from 32 participants.

**Table 1.**
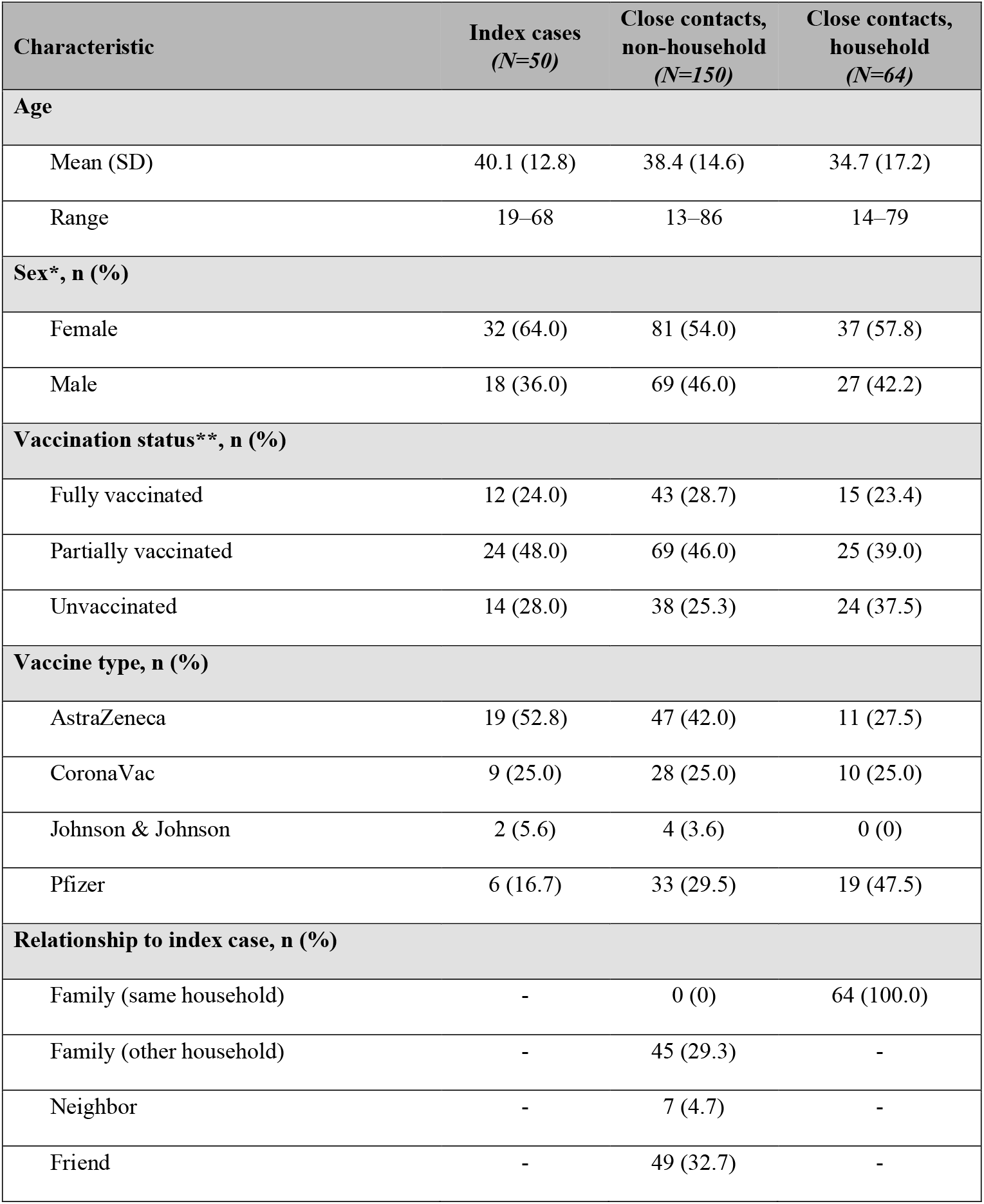
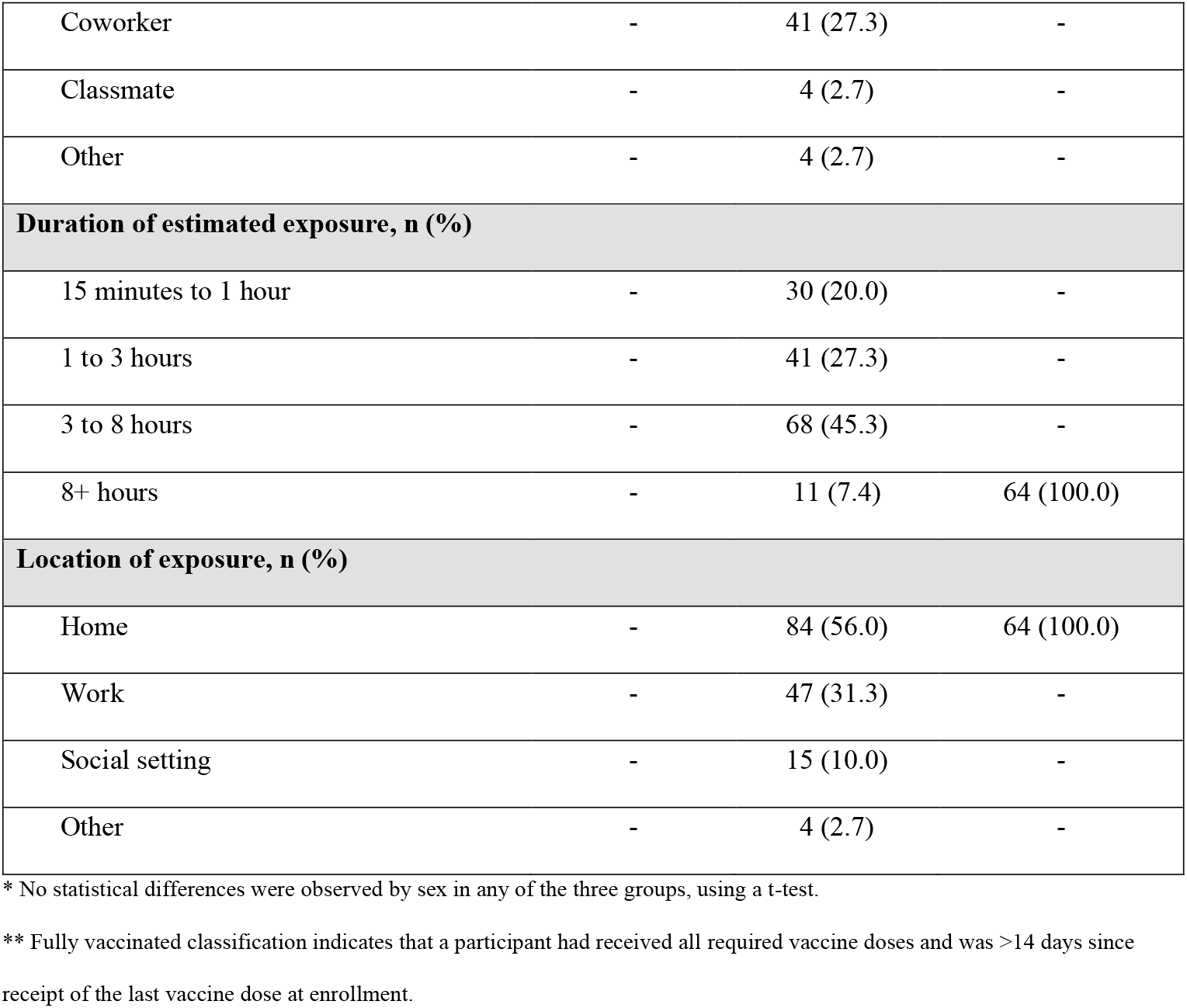
Characteristics of study participants.

| Characteristic | Index cases<br>(N=50) | Close contacts,<br>non-household<br>(N=150) | Close contacts,<br>household<br>(N=64) |
| --- | --- | --- | --- |
| **Age** |  |  |  |
| Mean (SD) | 40.1 (12.8) | 38.4 (14.6) | 34.7 (17.2) |
| Range | 19–68 | 13–86 | 14–79 |
| **Sex\*, n (%)** |  |  |  |
| Female | 32 (64.0) | 81 (54.0) | 37 (57.8) |
| Male | 18 (36.0) | 69 (46.0) | 27 (42.2) |
| **Vaccination status\*\*, n (%)** |  |  |  |
| Fully vaccinated | 12 (24.0) | 43 (28.7) | 15 (23.4) |
| Partially vaccinated | 24 (48.0) | 69 (46.0) | 25 (39.0) |
| Unvaccinated | 14 (28.0) | 38 (25.3) | 24 (37.5) |
| **Vaccine type, n (%)** |  |  |  |
| AstraZeneca | 19 (52.8) | 47 (42.0) | 11 (27.5) |
| CoronaVac | 9 (25.0) | 28 (25.0) | 10 (25.0) |
| Johnson & Johnson | 2 (5.6) | 4 (3.6) | 0 (0) |
| Pfizer | 6 (16.7) | 33 (29.5) | 19 (47.5) |
| **Relationship to index case, n (%)** |  |  |  |
| Family (same household) | - | 0 (0) | 64 (100.0) |
| Family (other household) | - | 45 (29.3) | - |
| Neighbor | - | 7 (4.7) | - |
| Friend | - | 49 (32.7) | - |
| Coworker | - | 41 (27.3) | - |
| Classmate | - | 4 (2.7) | - |
| Other | - | 4 (2.7) | - |
| **Duration of estimated exposure, n (%)** |  |  |  |
| 15 minutes to 1 hour | - | 30 (20.0) | - |
| 1 to 3 hours | - | 41 (27.3) | - |
| 3 to 8 hours | - | 68 (45.3) | - |
| 8+ hours | - | 11 (7.4) | 64 (100.0) |
| **Location of exposure, n (%)** |  |  |  |
| Home | - | 84 (56.0) | 64 (100.0) |
| Work | - | 47 (31.3) | - |
| Social setting | - | 15 (10.0) | - |
| Other | - | 4 (2.7) | - |

\* No statistical differences were observed by sex in any of the three groups, using a t-test.
\*\* Fully vaccinated classification indicates that a participant had received all required vaccine doses and was >14 days since receipt of the last vaccine dose at enrollment.

**Figure 1.**
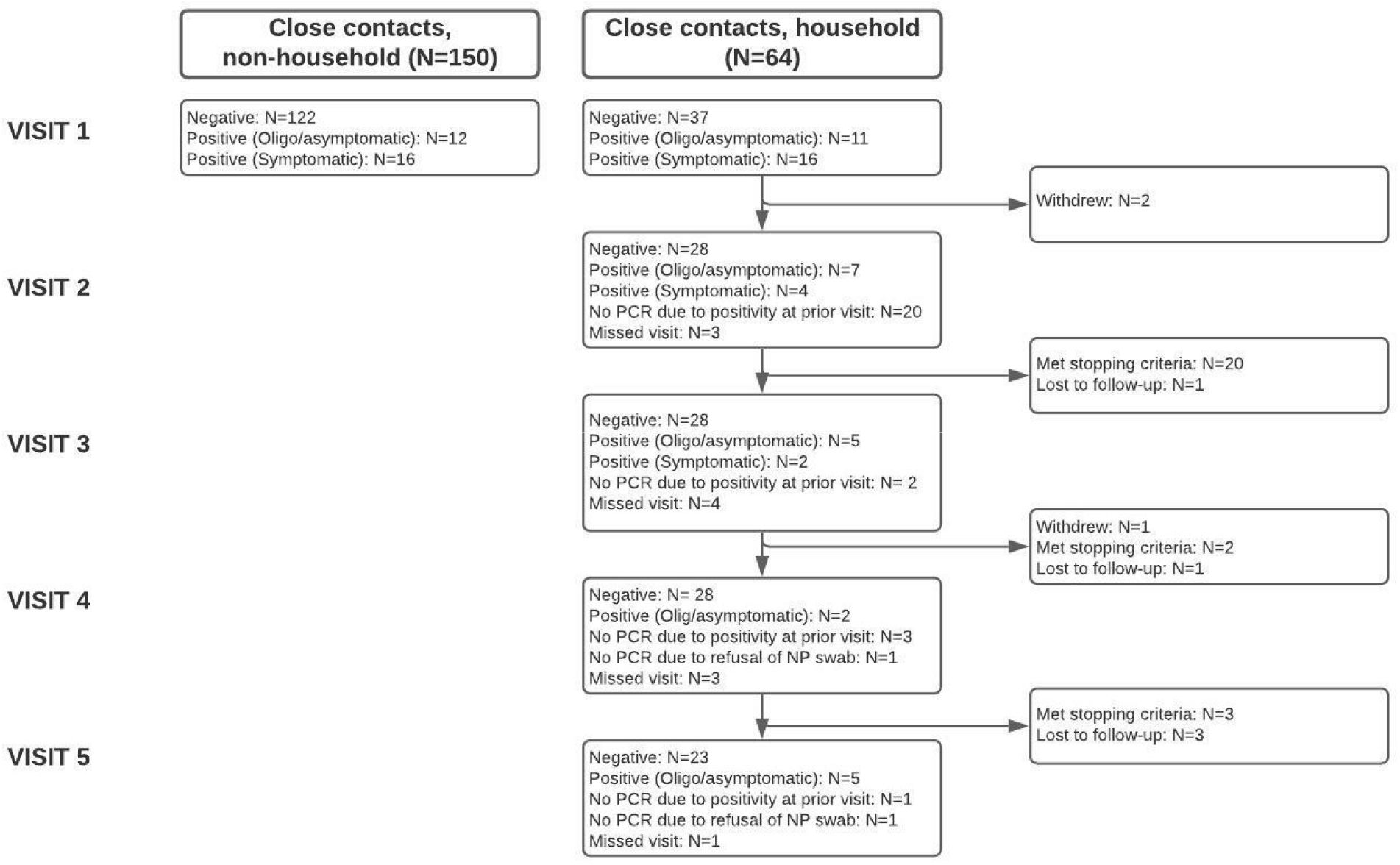
Status of study participants, by visit.

#### Vaccination status

Most participants were either partially (45%, 118/264) or fully (27%, 70/264) vaccinated at enrollment (Table 1). No statistical difference was observed in viral loads between vaccinated and unvaccinated individuals (Figure 2; Supplementary Material F, G).

**Figure 2.**
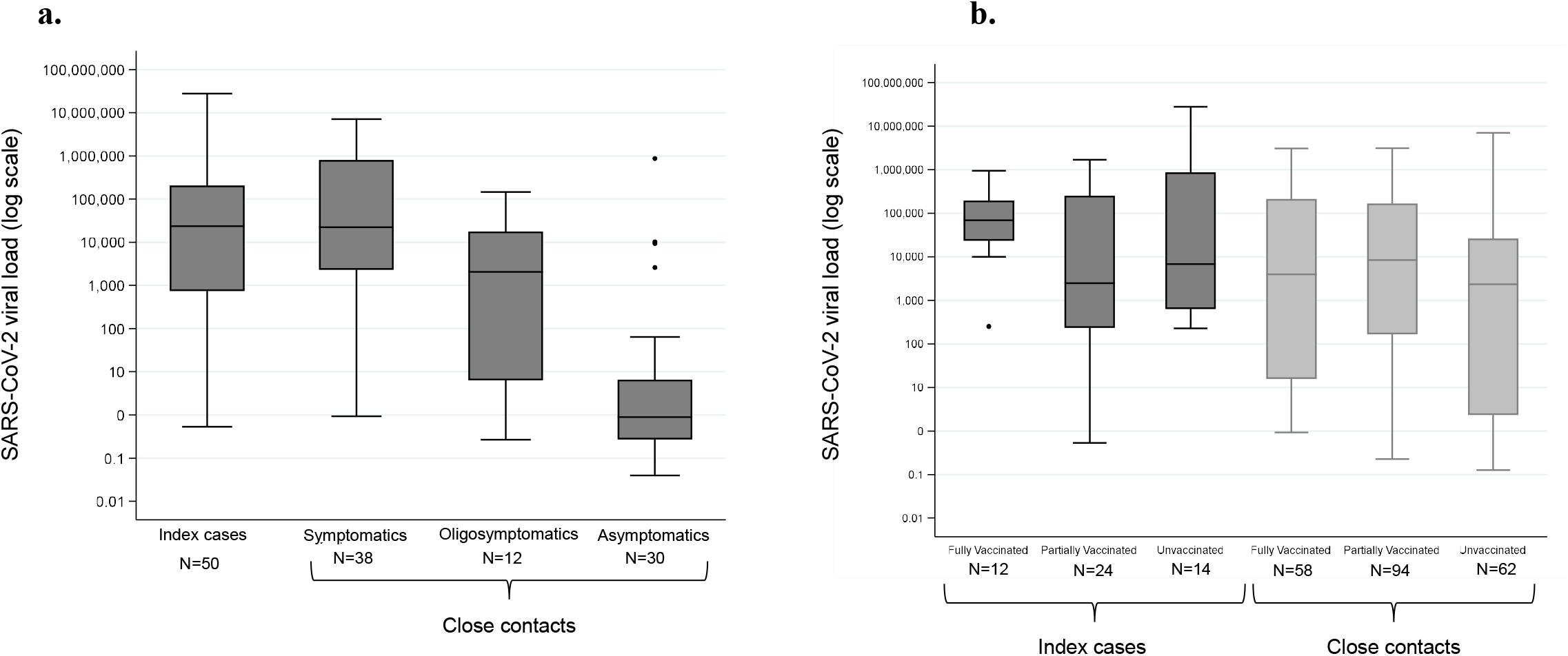
Viral load value relationships of study participants a) by infection category and b) by vaccination status.

#### Sequencing

Sequences were available for 84 positive samples: 68 Gamma (P.1, P.1.4, and P.1.7), and 16 Delta (AY.36, AY.4, AY.43, AY.99.2), with seven total lineages. The Delta strain became more prevalent among samples collected later in the study (Supplementary Material H).

### Diagnostic performance

The two POC ANS antigen tests demonstrated comparable performance, with overall sensitivity of 55.0% for the STANDARD Q (95% CI 43.5%–66.2%) and 50.6% for LumiraDx (95% CI 39.1%–62.1%) (Table 2). Performance increased to >80% sensitivity for both tests among symptomatic cases but decreased to <30% among oligo/asymptomatic cases. For specimens with Ct values less than 34, above which viral viability is negligent and quantification is not as reliable [34,35], performance of both tests improved, with sensitivities in the ranges of 90% and 60% for symptomatic and oligo/asymptomatic cases, respectively.

**Table 2.**
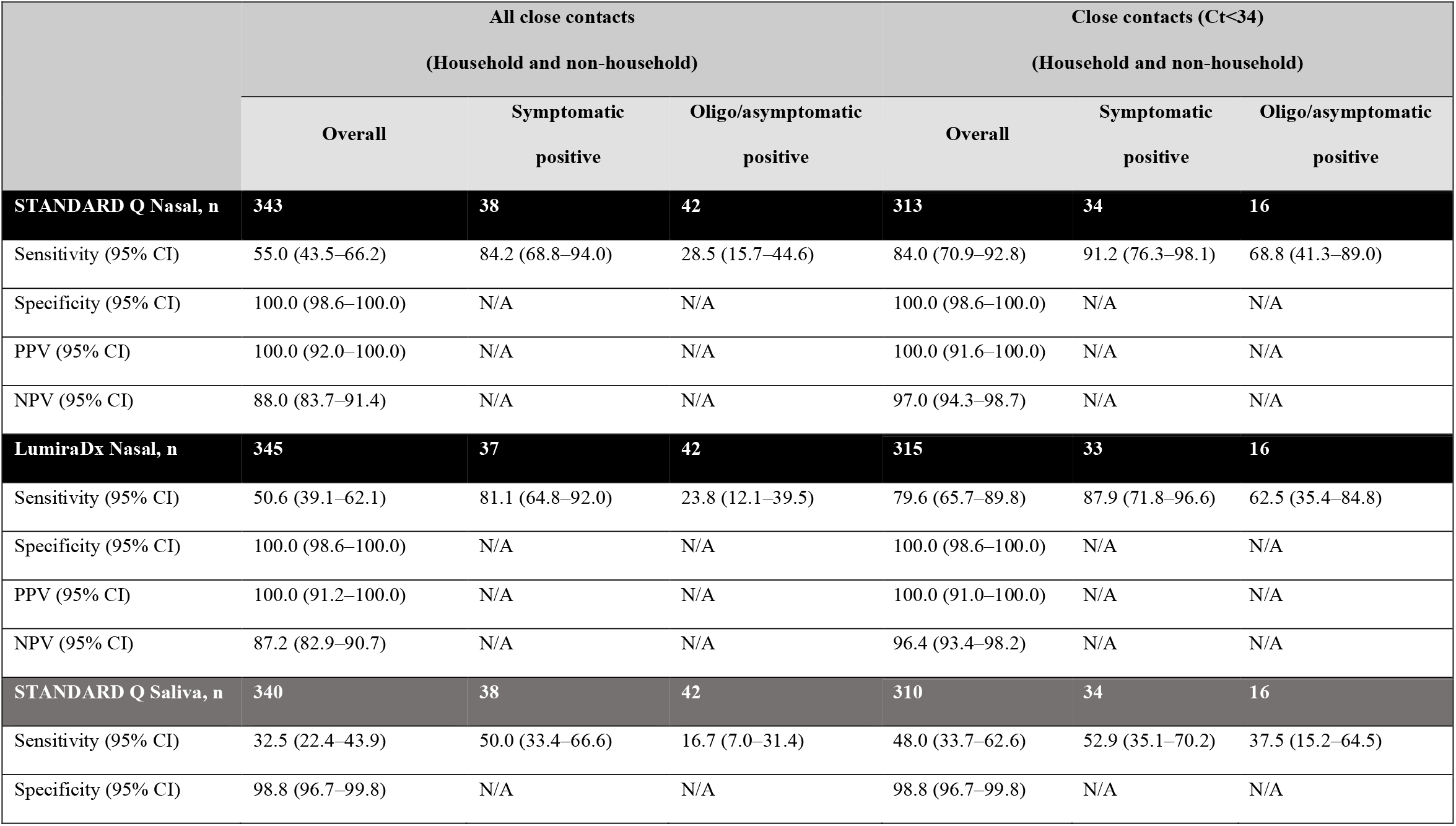
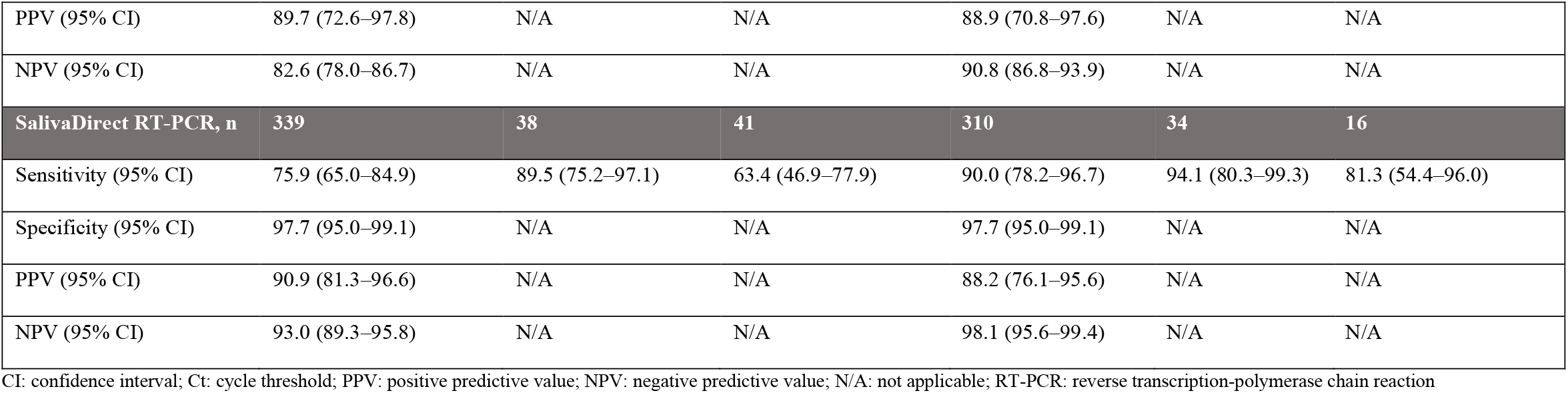
Performance indicators for tests evaluated using nasopharyngeal RT-PCR as the reference standard. Performance is shown across all close contacts and those with PCR Ct values of less than 34. Tests with black headers were run on anterior nares swabs, and tests with grey headers were run on saliva.

|  | All close contacts<br>(Household and non-household) |  |  | Close contacts (Ct<34)<br>(Household and non-household) |  |  |
| --- | --- | --- | --- | --- | --- | --- |
|  | Overall | Symptomatic<br>positive | Oligo/asymptomatic<br>positive | Overall | Symptomatic<br>positive | Oligo/asymptomatic<br>positive |
| **STANDARD Q Nasal, n** | **343** | **38** | **42** | **313** | **34** | **16** |
| Sensitivity (95% CI) | 55.0 (43.5–66.2) | 84.2 (68.8–94.0) | 28.5 (15.7–44.6) | 84.0 (70.9–92.8) | 91.2 (76.3–98.1) | 68.8 (41.3–89.0) |
| Specificity (95% CI) | 100.0 (98.6–100.0) | N/A | N/A | 100.0 (98.6–100.0) | N/A | N/A |
| PPV (95% CI) | 100.0 (92.0–100.0) | N/A | N/A | 100.0 (91.6–100.0) | N/A | N/A |
| NPV (95% CI) | 88.0 (83.7–91.4) | N/A | N/A | 97.0 (94.3–98.7) | N/A | N/A |
| **LumiraDx Nasal, n** | **345** | **37** | **42** | **315** | **33** | **16** |
| Sensitivity (95% CI) | 50.6 (39.1–62.1) | 81.1 (64.8–92.0) | 23.8 (12.1–39.5) | 79.6 (65.7–89.8) | 87.9 (71.8–96.6) | 62.5 (35.4–84.8) |
| Specificity (95% CI) | 100.0 (98.6–100.0) | N/A | N/A | 100.0 (98.6–100.0) | N/A | N/A |
| PPV (95% CI) | 100.0 (91.2–100.0) | N/A | N/A | 100.0 (91.0–100.0) | N/A | N/A |
| NPV (95% CI) | 87.2 (82.9–90.7) | N/A | N/A | 96.4 (93.4–98.2) | N/A | N/A |
| **STANDARD Q Saliva, n** | **340** | **38** | **42** | **310** | **34** | **16** |
| Sensitivity (95% CI) | 32.5 (22.4–43.9) | 50.0 (33.4–66.6) | 16.7 (7.0–31.4) | 48.0 (33.7–62.6) | 52.9 (35.1–70.2) | 37.5 (15.2–64.5) |
| Specificity (95% CI) | 98.8 (96.7–99.8) | N/A | N/A | 98.8 (96.7–99.8) | N/A | N/A |
| PPV (95% CI) | 89.7 (72.6–97.8) | N/A | N/A | 88.9 (70.8–97.6) | N/A | N/A |
| NPV (95% CI) | 82.6 (78.0–86.7) | N/A | N/A | 90.8 (86.8–93.9) | N/A | N/A |
| SalivaDirect RT-PCR, n | 339 | 38 | 41 | 310 | 34 | 16 |
| Sensitivity (95% CI) | 75.9 (65.0–84.9) | 89.5 (75.2–97.1) | 63.4 (46.9–77.9) | 90.0 (78.2–96.7) | 94.1 (80.3–99.3) | 81.3 (54.4–96.0) |
| Specificity (95% CI) | 97.7 (95.0–99.1) | N/A | N/A | 97.7 (95.0–99.1) | N/A | N/A |
| PPV (95% CI) | 90.9 (81.3–96.6) | N/A | N/A | 88.2 (76.1–95.6) | N/A | N/A |
| NPV (95% CI) | 93.0 (89.3–95.8) | N/A | N/A | 98.1 (95.6–99.4) | N/A | N/A |

CI: confidence interval; Ct: cycle threshold; PPV: positive predictive value; NPV: negative predictive value; N/A: not applicable; RT-PCR: reverse transcription-polymerase chain reaction

The SalivaDirect PCR assay showed the highest overall performance at 75.9% sensitivity (95% CI 65.0%–84.9%), which increased to 90.0% (95% CI 78.2%–96.7%) among contacts with Ct<34. In all scenarios, the rapid STANDARD Q Saliva Test had a sensitivity of <53%.

Figure 3 presents the viral load of positive specimens, stratified by results of the STANDARD Q Nasal and Saliva tests. Overall, specimens with low viral loads were more likely to yield negative results; however, misclassification of specimens with high viral loads was more common with the saliva test.

**Figure 3.**
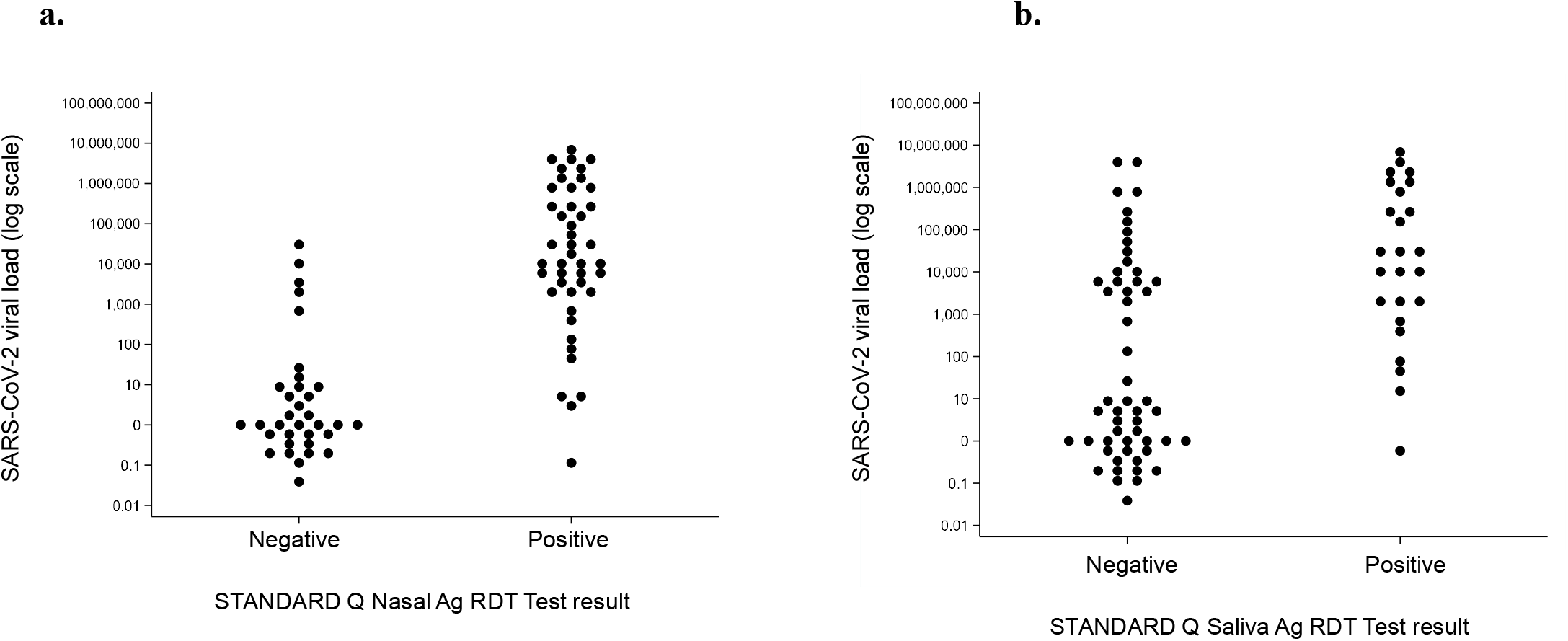
Viral load value distributions across antigen tests among close contacts for a) the STANDARD Q Nasal Test and b) the STANDARD Q Saliva Test.

### Longitudinal analysis

To investigate how test results changed over time, descriptive grid plots were generated for all household contacts with a positive reference result at any timepoint (Supplementary Material I). Figure 4 includes two examples of overall patterns in the dataset.

**Figure 4.**
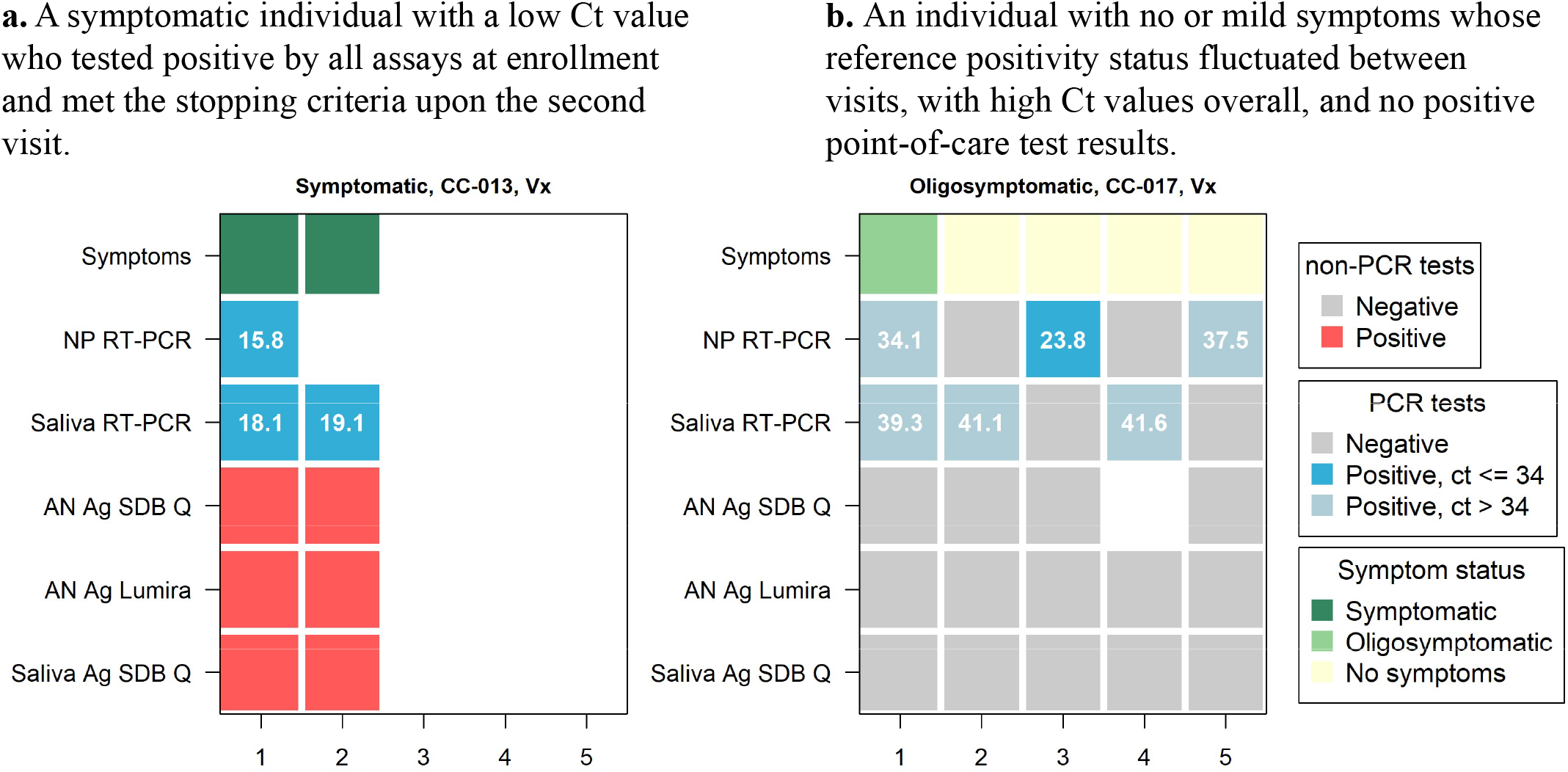
Descriptive plots for a subset of close contacts positive by the RT-PCR reference assay. Visit numbers are shown on the x-axis, and test results and symptom status are shown on the y-axis. Symptom status is presented independently of RT-PCR reference assay result.

The time-to-positivity from days since enrollment for close contacts with a positive reference result (Ct<34) at any timepoint was assessed by comparing the proportion of participants with positive results by reference RT-PCR and a POC ANS antigen test (STANDARD Q Nasal) under different scenarios for RT-PCR result turnaround time (Figure 5). Even with a relatively rapid RT-PCR result turnaround of 24 hours, >70% of contacts would have been identified by a POC test. At 48 hours, cumulative sensitivity is 80%, increasing to nearly 90% at four days.

**Figure 5.**
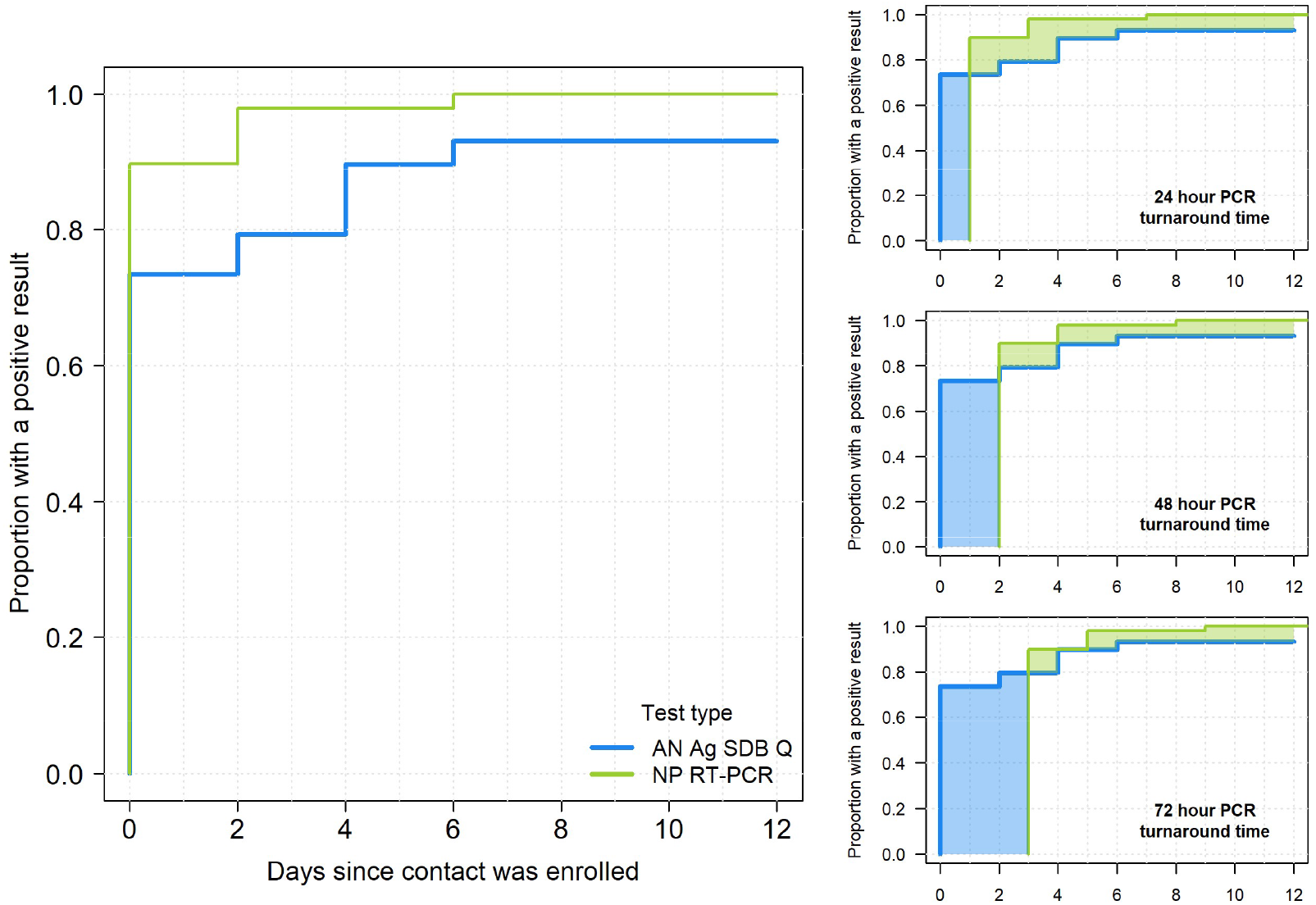
Time to positivity from time of first visit for close contacts with a positive NPS RT-PCR result (Ct<34) at any time. The blue line represents the proportion of NPS RT-PCR positive cases identified as positive by the point-of-care antigen test (STANDARD Q COVID-19 Ag test) on nasal samples, and the green line represents those identified by the reference NPS RT-PCR. Four different scenarios for RT-PCR result turnaround times are presented.

### Usability

In total, 12 study staff completed the usability assessment. All three POC antigen tests were considered easy to use and SUS scores were acceptable (>77) (Supplementary Material J).

## Discussion

In this study, performances of three POC antigen tests (two ANS and one saliva) and one molecular saliva assay were assessed among close contacts of COVID-19-positive index cases. All tests demonstrated strongest performance among symptomatic cases—and particularly those with Ct<34. Performance decreased among oligo/asymptomatic cases, which is consistent with results of prior studies [11, 13] and may indicate that the tests are best able to detect those most likely to be infectious [34, 35]. However, there is no universal Ct value cut-off-point that corresponds to infectivity, and the relationship between Ct values and viral load varies by laboratory [11].

The SalivaDirect assay had the best performance, with sensitivity of up to 90% among contacts with Ct<34. Although this assay uses a noninvasive sample type and a simplified procedure that minimizes processing time and costs, infrastructure and training requirements still limit the feasibility of implementing this test in many settings, with potential implications for time-to-results.

The saliva antigen test had the lowest overall performance, consistent with the findings of other evaluations of POC saliva antigen tests [36,37]. One recent evaluation of this test reported an overall sensitivity of 66.1%; however, the reference assay was conducted on saliva [38]. In this study, the test was run on passively collected saliva. This may have impacted performance, as the manufacturer recommends use of actively collected saliva with snorted nasal mucus.

The two POC ANS antigen tests—STANDARD Q Nasal and LumiraDx—demonstrated comparable performance which was best among cases with Ct<34. Among symptomatic cases and those with Ct<34, both tests met WHO performance criteria (≥ 80% sensitivity and ≥ 97% specificity) [10]. Both tests were also considered easy to use; however, the LumiraDx test requires use of an instrument.

Overall, the observed positivity rate among close contacts (65/214, 30%) highlights the importance of contact tracing and testing as a public health strategy [39]. The longitudinal data demonstrate the value of serial testing (particularly for individuals with known exposures) and the practical benefits of timely results [40, 41]. In this study, we show that in settings where RT-PCR is unavailable or where time-to-results is >4 days, close to 90% of individuals with Ct<34 could benefit from an earlier result via a POC test. Even in settings where RT-PCR results are available within 24 hours, cumulative sensitivity of a POC test is >70%. With repeat serial testing over a period of 9 days, the cumulative sensitivity of a POC ANS antigen test increases from 70% to near 90%. In many settings, limited RT-PCR testing capacity— especially during high demand—can lead to delays in results. Immediate results can impact behavior of potentially infectious individuals, encouraging earlier isolation and signaling where additional testing is warranted [4]. The emergence of antiviral therapies—which are more effective the sooner they are taken—further underscores the value of timely results.

## Data Availability

All data included in the present work are available online at https://dataverse.harvard.edu/dataset.xhtml?persistentId=doi:10.7910/DVN/NGNUXY (doi:10.7910/DVN/NGNUXY).

## Limitations

Limitations of the study include its modest sample size, reflected in the 95% Cis reported with performance indicators. Further, the STANDARD Q Nasal and LumiraDx tests are among the best-in-class commercial POC antigen tests. Other tests with lower performance may increase the risk of missing infections against the benefit of identifying cases, to the extent that other strategies may be needed if RT-PCR is unavailable. Lastly, future research should investigate implications of new variants on diagnostic performance across sample types.

## Conclusion

The near immediate time-to-result of rapid antigen tests is a significant benefit that offsets reduced sensitivity by decreasing diagnostic delays and onward viral transmission. Here, we demonstrate that POC ANS antigen tests for SARS-CoV-2 are easy to use and perform adequately to provide prompt, actionable information to both the health system and individuals.

## Funding

This work was supported by grants from The Rockefeller Foundation [2020 HTH 039] and Amazon.com [2D-04020007] to GJD. REDCap hosted at the Institute of Translational Health Sciences is supported by the National Center For Advancing Translational Sciences of the National Institutes of Health under Award Number UL1 TR002319. FGN and VAN were supported by the National Council for Scientific and Technological Development [grant 403276/2020-9] and Inova Fiocruz/Fundação Oswaldo Cruz [grant VPPCB-007-FIO-18-2-30 - Knowledge generation]. FGN is a CNPq fellow.

## Acknowledgements

The authors would like to thank all study participants as well as the clinical and laboratory staff at CEPEM involved with this study. We also thank SD Biosensor and LumiraDx for facilitating the availability of their tests for this study. Finally, we also thank Amanda Tsang and Christine Waresak for editorial support with the manuscript.

## Notes

*Conflict of Interest Statement* The authors declare no conflicts of interest.

### Competing Interest Statement

The authors have declared no competing interest.

### Author Declarations

WCG Institutional Review Board gave ethical approval for this work (1301165). The ethics committee of the Centro de Pesquisa em Medicina Tropical de Rondonia (CEPEM) and National Research Ethics Commission of Brazil also approved this study (44351421.0.0000.0011).

## References

1. Ministry of Health Health Surveillance Secretariat. Special Epidemiological Bulletin: Coronavirus Disease COVID-19. 10 December 2021. Available at: https://www.gov.br/saude/pt-br/media/pdf/2021/dezembro/11/boletim_epidemiologico_covid_92_10dez21.pdf.

2. Vandenberg O, Martiny D, Rochas O, van Belkum A, Kozlakidis Z. Considerations for diagnostic COVID-19 tests. Nat Rev Microbiol 2021 ; 19(3):171–183. doi:10.1038/s41579-020-00461-z.

3. Kretzschmar ME, Rozhnova G, Bootsma MCJ, van Boven M, van de Wijgert JHHM, Bonten MJM. Impact of delays on effectiveness of contact tracing strategies for COVID-19: a modelling study. Lancet Public Health 2020 ; 5(8):e452–e459. doi:10.1016/S2468-2667(20)30157-2.

4. Larremore DB, Wilder B, Lester E, et al. Test sensitivity is secondary to frequency and turnaround time for COVID-19 screening. Sci Adv 2021 ; 7(1):eabd5393. doi:10.1126/sciadv.abd5393.

5. Sah P, Fitzpatrick MC, Zimmer CF, et al. Asymptomatic SARS-CoV-2 infection: a systematic review and meta-analysis. Proc Natl Acad Sci U S A 2021 ; 118(34):e2109229118. doi:10.1073/pnas.2109229118.

6. Buitrago-Garcia D, Egli-Gany D, Counotte MJ, et al. Occurrence and transmission potential of asymptomatic and presymptomatic SARS-CoV-2 infections: a living systematic review and meta-analysis. PLoS Med 2020 ; 17(9):e1003346. doi:10.1371/journal.pmed.1003346.

7. Byambasuren O, Cardona M, Bell K, et al. Estimating the extent of asymptomatic COVID-19 and its potential for community transmission: Systematic review and meta-analysis. JAMMI 2020 ; 5(4): 223–234. doi: 10.3138/jammi-2020-0030.

8. Qiu X, Nergiz AI, Maraolo AE, et al. The role of asymptomatic and pre-symptomatic infection in SARS-CoV-2 transmission-a living systematic review. Clin Microbiol Infect 2021 ; 27(4): 511– 519. doi: 10.1016/j.cmi.2021.01.011.

9. Fiore VG, DeFelice N, Glicksberg BS, et al. Containment of COVID-19: Simulating the impact of different policies and testing capacities for contact tracing, testing, and isolation. PLoS One 2021 ; 16(3):e0247614. doi:10.1371/journal.pone.0247614.

10. World Health Organization. Antigen-detection in the diagnosis of SARS-CoV-2 infection. Interim Guidance. 6 October 2021. Geneva; WHO. Available at: https://www.who.int/publications/i/item/antigen-detection-in-the-diagnosis-of-sars-cov-2infection-using-rapid-immunoassays.

11. Dinnes J, Deeks JJ, Adriano A, et al. Rapid, point-of-care antigen and molecular-based tests for diagnosis of SARS-CoV-2 infection. Cochrane Database Syst Rev 2020 ; 8(8):CD013705. doi:10.1002/14651858.CD013705.

12. Drain P. Rapid diagnostic testing for SARS-CoV-2 [published online ahead of print, 2022 Jan 7]. N Eng J Med 2022. doi:10.1056/NEJMcp2117115.

13. Brümmer LE, Katzenschlager S, Gaeddert M, et al. Accuracy of novel antigen rapid diagnostics for SARS-CoV-2: a living systematic review and meta-analysis [published correction appears in PLoS Med 2021 Oct 13 ; 18(10):e1003825]. PLoS Med 2021 ; 18(8):e1003735. doi:10.1371/journal.pmed.1003735.

14. Boum Y, Fai KN, Nikolay B, et al. Performance and operational feasibility of antigen and antibody rapid diagnostic tests for COVID-19 in symptomatic and asymptomatic patients in Cameroon: a clinical, prospective, diagnostic accuracy study [published correction appears in Lancet Infect Dis 2021 Jun ; 21(6):e148] [published correction appears in Lancet Infect Dis. 2021 Jul ; 21i7):e182]. Lancet Infect Dis 2021 ; 21(8):1089–96. doi:10.1016/S1473-3099(21)00132-8.

15. Baro B, Rodo P, Ouchi D, et al. Performance characteristics of five antigen-detecting rapid diagnostic test (Ag-RDT) for SARS-CoV-2 asymptomatic infection: a head-to-head benchmark comparison. J Infect 2021 ; 82(6):269 –275. doi: 10.1016/j.jinf.2021.04.009

16. Torres I, Poujois S, Albert E, Colomina J, Navarro D. Evaluation of a rapid antigen test (Panbio™ COVID-19 Ag rapid test device) for SARS-CoV-2 detection in asymptomatic close contacts of COVID-19 patients. Clin Microbiol Infect 2021 ; 27(4):636.e1-636.e4. doi:10.1016/j.cmi.2020.12.022.

17. Schuit E, Veldhuijzen IK, Venekamp RP, et al. Diagnostic accuracy of rapid antigen tests in asymptomatic and presymptomatic close contacts of individuals with confirmed SARS-CoV-2 infection: cross sectional study. BMJ 2021 ; 374:n1676. doi:10.1136/bmj.n1676.

18. Ministry of Health. Epidemiological Surveillance Guide: Public Health Emergency of National Importance due to Coronavirus Disease 2019. Available at: https://portalarquivos.saude.gov.br/images/af_gvs_coronavirus_6ago20_ajustes-finais-2.pdf.

19. Drain P, Sulaiman R, Hoppers M, Lindner NM, Lawson V, Ellis JE. Performance of the LumiraDx Microfluidic Immunofluorescence Point-of-Care SARS-CoV-2 Antigen Test in asymptomatic adults and children [published online ahead of print, 2021 Oct 20]. Am J Clin Pathol 2021 ; aqab173. doi:10.1093/ajcp/aqab173.

20. Kohmer N, Toptan T, Pallas C, et al. The comparative clinical performance of four SARS-CoV-2 rapid antigen tests and their correlation to infectivity in vitro. J Clin Med 2021 ; 10(2):328. doi:10.3390/jcm10020328.

21. Krüger LJ, Klein JAF, Tobian F, et al. Evaluation of accuracy, exclusivity, limit-of-detection and ease-of-use of LumiraDx™: An antigen-detecting point-of-care device for SARS-CoV-2 [published online ahead of print, 2021 Aug 12]. Infection 2021 ; 1–12. doi:10.1007/s15010-021-01681-y.

22. Bianco G, Boattini M, Barbui AM, et al. Evaluation of an antigen-based test for hospital point-of-care diagnosis of SARS-CoV-2 infection. J Clin Virol 2021 ; 139:104838. doi:10.1016/j.jcv.2021.104838.

23. Fekete T. In adults and children, a rapid POC antigen test for COVID-19 (LumiraDx) had ≥97% sensitivity and specificity vs. RT-PCR. Ann Intern Med 2021 ; 174(7):JC82. doi:10.7326/ACPJ202107200-082.

24. Karon BS, Donato LJ, Bridgeman AR, et al. Analytical sensitivity and specificity of four point of care rapid antigen diagnostic tests for SARS-CoV-2 using real-time quantitative PCR, quantitative droplet digital PCR, and a mass spectrometric antigen assay as comparator methods. Clin Chem 2021 ; 67(11):1545–1553. doi:10.1093/clinchem/hvab138.

25. Vogels CBF, Watkins AE, Harden CA, et al. SalivaDirect: A simplified and flexible platform to enhance SARS-CoV-2 testing capacity. Med (N Y) 2021 ; 2(3):263-280.e6. doi:10.1016/j.medj.2020.12.010.

26. Rodríguez Flores SN, Rodríguez-Martínez LM, Reyes-Berrones BL, Fernández-Santos NA, Sierra-Moncada EJ, Rodríguez-Pérez MA. Comparison between a standard and SalivaDirect RNA extraction protocol for molecular diagnosis of SARS-CoV-2 using nasopharyngeal swab and saliva clinical samples. Front Bioeng Biotechnol 2021 ; 9:638902. doi:10.3389/fbioe.2021.638902.

27. Freppel W, Merindol N, Rallu F, Bergevin M. Efficient SARS-CoV-2 detection in unextracted oro-nasopharyngeal specimens by rRT-PCR with the Seegene Allplex™ 2019-nCoV assay. Virol J 2020 ; 17(1):196. doi: 10.1186/s12985-020-01468-x.

28. World Health Organization. Antigen detection rapid diagnostic tests for coronavirus disease 2019 (COVID-19): master protocol for monitored implementation. Version 1.0. Geneva: WHO, 25 November 2020. Available at: https://www.who.int/news-room/articles-detail/sars-cov-2-antigen-detecting-rapid-diagnostic-test-implementation-proposals.

29. Sauro, J. A practical guide to the System Usability Scale: Background, benchmarks, & best practices. Denver, CO: Measuring Usability LLC, 2011.

30. Bangor, A., Kortum, PT, & Miller, JT. (2008). An empirical evaluation of the System Usability Scale. International Journal of Human-Computer Interaction 2008 ; 24(6), 574–94. https://doi.org/10.1080/10447310802205776.

31. United States Food and Drug Administration. Template for Developers of Antigen Tests. Available at: https://www.fda.gov/medical-devices/coronavirus-disease-2019-covid-19-emergency-use-authorizations-medical-devices/in-vitro-diagnostics-euas. Version October 26 2020. Accessed 24 November 2020.

32. Centers for Disease Control and Prevention. Coronavirus Disease 2019 (COVID-19) 2020 Interim Case Definition. Available at: https://ndc.services.cdc.gov/case-definitions/coronavirus-disease-2019-2020/. Accessed 10 September 2021.

33. Harris PA, Taylor R, Thielke R, Payne J, Gonzalez N, Conde JG. Research electronic data capture (REDCap)—a metadata-driven methodology and workflow process for providing translational research informatics support, J Biomed Inform 2009 ; Apr;42(2):377–81.

34. Routsias JG, Mavrouli M, Tsoplou P, Dioikitopoulou K, Tsakris A. Diagnostic performance of rapid antigen tests (RATs) for SARS-CoV-2 and their efficacy in monitoring the infectiousness of COVID-19 patients. Sci Rep 2021 ; 11:22863.

35. La Scola B, Le Bideau M, Andreani J, et al. Viral RNA load as determined by cell culture as a management tool for discharge of SARS-CoV-2 patients from infectious disease wards. Eur J Clin Microbiol Infect Dis 2020 ; 39:1059–61.

36. Yokota I, Sakurazawa T, Sugita J, et al. Performance of qualitative and quantitative antigen tests for SARS-CoV-2 using saliva. Infect Dis Rep 2021 ; 13(3):742–7. doi:10.3390/idr13030069.

37. Kritikos A, Caruana G, Brouillet R, et al. Sensitivity of rapid antigen testing and RT-PCR performed on nasopharyngeal swabs versus saliva samples in COVID-19 hospitalized patients: results of a prospective comparative trial (RESTART). Microorganisms 2021 ; 9(9):1910. doi:10.3390/microorganisms9091910.

38. Igloi Z, Velzing J, Huisman R, et al. Clinical evaluation of the SD Biosensor saliva antigen rapid test with symptomatic and asymptomatic, non-hospitalized patients. PLoS One 2021 ; Dec 22;16(12):e0260894. doi:10.1371/journal.pone.0260894.

39. Chung S, Marlow S, Tobias N, et al. Lessons from countries implementing find, test, trace, isolation and support policies in the rapid response of the COVID-19 pandemic: a systematic review. BMJ Open 2021 ; 11(7):e047832. Doi: 10.1136.bmjopen-2020-047832.

40. Smith RL, Gibson LL, Martinez PP, et al. Longitudinal assessment of diagnostic test performance over the course of acute SARS-CoV-2 infection. J Infect Dis 2021 ; 224(6). doi:10.1093/infdis/jiab337.

41. Revollo B, Blanco I, Soler P, et al. Same-day SARS-CoV-2 antigen test screening in an indoor mass-gathering live music event: a randomized controlled trial. Lancet Infect Dis 2021 ; 21(10):P1365 – 1372. doi: 10.1016/S1473-3099(21)00268-1.

