## Supplemental Information for "Screening for SARS-CoV-2 in close contacts of individuals with confirmed infection: performance and operational considerations"

**Supplementary Material**

**Supplementary Material A.** Detailed description of inclusion/exclusion criteria, recruitment, and enrollment methods.

Participants in this study included both symptomatic index cases with confirmed COVID-19, as well their close contacts. Eligible index cases were recruited through existing clinical care platforms in Porto Velho. Close contacts were identified through a contact elicitation interview that was administered to the index cases at enrollment. Specific inclusion/exclusion criteria and recruitment/enrollment procedures for these two populations are described below.

Inclusion criteria

- Index cases
  - 18 years of age or older.
  - Positive screening result for SARS-CoV-2 infection according to an Anvisa-approved antigen rapid diagnostic test.
  - Presentation of an acute respiratory condition with at least two of the following signs and symptoms consistent with the clinical case definition for COVID-19 in Brazil: fever, chills, sore throat, headache, cough, runny nose, or olfactory/taste disorder(s).
  - Within 7 days of symptom onset.
  - Willing and able to provide informed consent to participate and comply with study requirements.
- Close contacts
  - Close contact with an enrolled study index case, defined as meeting one or more of the following criteria within the investigation period of the index case (2 days prior to symptom onset to the time of the interview):
    - Less than 1 meter away from an index case for a minimum of 15 minutes.
    - Had direct physical contact (e.g., shaking hands) with an index case.
    - Is a health professional who provided care to the index case without using appropriate personal protective equipment (PPE).
  - 12 years of age or older.
  - Resides within the municipality of Porto Velho.
  - Willing and able to provide informed consent and comply with the study requirements.

Exclusion criteria

- Index cases
  - Any study site employees who are involved in the protocol or may have access to study-related data.
  - Unwilling or unable to provide informed consent and/or comply with study requirements.
- Close contacts
  - Prior positive test result on a COVID-19 test of any type within the past 3 months (self-report).
  - Any study site employees who are involved in the protocol or may have access to study-related data.
  - Unwilling or unable to provide informed consent and/or comply with study requirements.

Recruitment and enrollment: Index cases

Index cases were recruited through existing clinical care platforms in Porto Velho. Patients and health workers from collaborating facilities and outpatient clinics who were presenting signs and symptoms consistent with COVID-19 were pre-screened against the eligibility criteria. Individuals who were within 7 days of symptom onset, 18 years of age or older, and able to participate were invited to participate in the study. If the person expressed interest, he or she completed the informed consent process. Next, all participants were screened with an Anvisa-approved rapid antigen test for SARS-CoV-2 at the point of care (the STANDARD Q COVID-19 Ag Nasal Test). All participants with negative results on the screening test were considered screen failures and exited the study. All participants with positive screening results were enrolled into the study, completed the remaining study procedures (including completion of the contact elicitation interview), and were referred to the public health system for clinical care.

Recruitment and enrollment: Close contacts

Following identification of SARS-CoV-2 infection in an index case according to the rapid screening test, all associated close contacts identified during the subject’s contact elicitation interview were notified of their exposure by phone within 72 hours. All close contacts, regardless of their participation in the study, received standard information in accordance with national and municipal guidelines for contact tracing that included information about their potential exposure, instruction on recommended isolation procedures, information on when/how they should seek care, and information on resources available for their care and testing from the municipality. For eligible close contacts who expressed interest in participating, study staff scheduled a time for a home visit where potential participants were again screened against the enrollment criteria, completed the informed consent process, and were enrolled into the study.

**Supplementary Material B.** Detailed description of specimen collection and management methods.

Specimen collection

First, two paired anterior nares swabs (ANS) were collected simultaneously using the swabs included in the STANDARD Q COVID-19 Ag Nasal and LumiraDx test kits. Both nostrils were swabbed using each of the two swabs: each swab was used first in one nostril as the “first pass” swab, before they were switched and used with the opposite nostril as the “second-pass” swab. The STANDARD Q COVID-19 Ag Nasal Test swab was then placed into the buffer solution associated with that test, which was performed on-site at the point of care. The LumiraDx swab was added to the test’s extraction buffer solution and transferred to a cryovial for transport back to the laboratory. Next, the nasopharyngeal swab (NPS) (Firstlab, Jiangsu Rongye Technology, China) was collected and placed into 3 mL of viral transport media (Laborclin, Brazil). Finally, a minimum of 0.5 mL of saliva was collected into a sterile container. Saliva was collected passively from saliva that pools naturally in the mouth; participants were not asked to cough or sniff prior to specimen collection. Participants did not ingest any water in the 10 minutes prior to collection and did not smoke or consume any other drinks, food, or nasal sprays for 30 minutes prior to collection.

Specimen transport

All specimens were transported from the visit site (participant’s home or clinical facility) to the CEPEM laboratory in cold storage (2ºC–8ºC) within 4 hours of collection.

**Supplementary Material C.** Detailed description of genomic sequencing methods.

Genomic sequencing for this study was conducted using Illumina COVIDSEQ with some modifications as previously described by Naveca et al. [1], on MiSeq or NextSeq 1000 sequencers. Reads were assembled using DRAGEN COVID Lineage 3.5.4 at Illumina BaseSpace (http://basespace.illumina.com) or BBMap 38.84 embedded in Geneious Prime 2022.0.1. Consensus sequences were analyzed for quality issues with Nextclade (<https://clades.nextstrain.org>) [2], and lineages were identified using the pangolin tool on its most up-to-date version [3].

All SARS-CoV-2 genomes generated and analyzed in this study are available at the EpiCoV database in GISAID (<https://www.gisaid.org>).

References:

1. Naveca FG, Nascimento V, de Souza VC, et al. COVID-19 in Amazonas, Brazil, was driven by the persistence of endemic lineages and P.1 emergence. Nat Med **2021** ; 27, 1230–1238. doi: 10.1038/s41591-021-01378-7
2. Askamentov I, Roemer C, Hodcroft EB, et al. Nextclade: clade assignment, mutation calling and quality control for viral genomes. Journal of Open Source Software **2021** ; 6(67), 3773. doi: 10.21105/joss.03773
3. O’Toole A, Scher E, Underwood A, et al. Assignment of epidemiological lineages in an emerging pandemic using the pangolin tool. Virus Evolution **2021** ; 7(2), veab064. doi: 10.1093/ve/veab064.

**Supplementary Material D.** Usability questionnaire.

USABILITY QUESTIONNAIRE

| Participant ID | U - ___ ___ |
| --- | --- |
| Date (YYYY/MM/DD) | __ __ __ __ / __ __ / __ __ |
| Test name *(select all that apply)* | ⬜ STANDARD Q COVID-19 Ag: Nasal  ⬜ STANDARD Q COVID-19 Ag: Saliva  ⬜ LumiraDx Ag SARS-CoV-2 Ag Test |

**BACKGROUND**

| Occupation | ⬜ Laboratory technician  ⬜ Medical doctor  ⬜ Registered nurse  ⬜ Field agent  ⬜ Other, specify: ____________ |
| --- | --- |
| Educational level | ⬜ High school (Ensino médio)  ⬜ Technical certificate (Ensino técnico)  ⬜ University, bachelors  ⬜ University, masters  ⬜ University, doctorate  ⬜ Medical school  ⬜ Other, specify: ___________  ⬜ Prefer not to answer |
| Years of relevant experience | ⬜ Less than 1  ⬜ 1 to 3  ⬜ 3 to 5  ⬜ 5 to 10  ⬜ More than 10  ⬜ Prefer not to answer |
| How many times have you performed this test? | ⬜ None  ⬜ 1 to 10  ⬜ 11 to 50  ⬜ 51 to 100  ⬜ >100 |

**SYSTEMS USABILITY SCALE**

1. **I would like to use this device frequently**

Strongly Disagree	Disagree	Neither Agree or Disagree	Agree	Strongly Agree

2. **I found this device unnecessarily complex**

Strongly Disagree	Disagree	Neither Agree or Disagree	Agree	Strongly Agree

3. **I thought the device was easy to use**

Strongly Disagree	Disagree	Neither Agree or Disagree	Agree	Strongly Agree

4. **I would need the support of a technical person to be able to use this device**

Strongly Disagree	Disagree	Neither Agree or Disagree	Agree	Strongly Agree

5. **I found that the various steps in this device were well integrated**

Strongly Disagree	Disagree	Neither Agree or Disagree	Agree	Strongly Agree

6. **I thought that there was too much inconsistency in this device**

Strongly Disagree	Disagree	Neither Agree or Disagree	Agree	Strongly Agree

7. **I would imagine that most people would learn to use this device very quickly**

Strongly Disagree	Disagree	Neither Agree or Disagree	Agree	Strongly Agree

8. **I found the device very awkward to use**

Strongly Disagree	Disagree	Neither Agree or Disagree	Agree	Strongly Agree

9. **I feel very confident using the device**

Strongly Disagree	Disagree	Neither Agree or Disagree	Agree	Strongly Agree

10. **I needed to learn a lot of things before I could use this device**

Strongly Disagree	Disagree	Neither Agree nor Disagree	Agree	Strongly Agree

**EASE OF USE QUESTIONNAIRE: STANDARD Q COVID-19 Ag**

| **Background** | | |
| --- | --- | --- |
| Which version of the test are you commenting on: | ⬜ Nasal  ⬜ Saliva | |
| **General questions** | | |
| 1. Overall, how satisfied are you with the test? | | Not at all Very  satisfied satisfied    1 2 3 4 5  ⬜ ⬜ ⬜ ⬜ ⬜ |
| 2. What do you like most about the test? | |  |
| 3. What do you like least about the test? | |  |
| **Kit components** | | |
| 4. How satisfied are you with each of the kit components? | | Not at all Very  satisfied satisfied    1 2 3 4 5 Don’t know |
| External paper box | | ⬜ ⬜ ⬜ ⬜ ⬜ ⬜ |
| Extraction buffer tube | | ⬜ ⬜ ⬜ ⬜ ⬜ ⬜ |
| Extraction buffer nozzle cap | | ⬜ ⬜ ⬜ ⬜ ⬜ ⬜ |
| Buffer tube rack | | ⬜ ⬜ ⬜ ⬜ ⬜ ⬜ |
| Swab for specimen collection | | ⬜ ⬜ ⬜ ⬜ ⬜ ⬜ |
| Test device | | ⬜ ⬜ ⬜ ⬜ ⬜ ⬜ |
| Test device foil pouch | | ⬜ ⬜ ⬜ ⬜ ⬜ ⬜ |
| Positive/negative control swabs | | ⬜ ⬜ ⬜ ⬜ ⬜ ⬜ |
| 5. Which kit components do you think could be improved? *Select all that apply* | | ⬜ External paper box of kit  ⬜ Extraction buffer tube  ⬜ Extraction buffer nozzle cap  ⬜ Buffer tube rack  ⬜ Swab for specimen collection  ⬜ Test device  ⬜ Test device foil pouch  ⬜ Positive/negative control swabs |
| 6. How would you improve these components? | |  |
| **Device design** | | |
| 7. Overall, how satisfied are you with the design of the device? | | Not at all Very  satisfied satisfied    1 2 3 4 5  ⬜ ⬜ ⬜ ⬜ ⬜ |
| 8. How satisfied are you with the design of the following features? | | Not at all Very  satisfied satisfied    1 2 3 4 5 |
| Size of test device | | ⬜ ⬜ ⬜ ⬜ ⬜ |
| Size of the well to add the extracted sample | | ⬜ ⬜ ⬜ ⬜ ⬜ |
| Size of reading window | | ⬜ ⬜ ⬜ ⬜ ⬜ |
| Logical sequence of steps | | ⬜ ⬜ ⬜ ⬜ ⬜ |
| 9. Which design features do you think could be improved? *Select all that apply* | | ⬜ Size of test device  ⬜ Size of the well to add sample mix  ⬜ Size of reading window  ⬜ Logical sequence of steps  ⬜ Other, specify:____________________ |
| 10. How would you improve these components? | |  |
| **Ease of use** | | |
| 11. On a scale from 1 to 5, please rate the ease of using this test. | | Not at all Very  easy to use easy to use    1 2 3 4 5  ⬜ ⬜ ⬜ ⬜ ⬜ |
| 12. On a scale of 1 to 5, how difficult were the following steps? | | Not at Very  all difficult difficult  1 2 3 4 5 |
| Check expiry date | | ⬜ ⬜ ⬜ ⬜ ⬜ |
| Remove the test device from the pouch | | ⬜ ⬜ ⬜ ⬜ ⬜ |
| Open the extraction buffer tube | | ⬜ ⬜ ⬜ ⬜ ⬜ |
| Insert the swab into the tube | | ⬜ ⬜ ⬜ ⬜ ⬜ |
| Ease of swab extraction procedure | | ⬜ ⬜ ⬜ ⬜ ⬜ |
| Ability to perform extraction procedure consistently | | ⬜ ⬜ ⬜ ⬜ ⬜ |
| Transfer the correct number of drops into the sample well of the test device | | ⬜ ⬜ ⬜ ⬜ ⬜ |
| 13. Have you had any challenges with using this test? | | ⬜ Yes  ⬜ No  If yes, please explain: |
| 14. Did you collect participant samples for this test? | | ⬜ Yes  ⬜ No |
| 15. *[If yes to #14]* Please rate your experience collecting samples for this test. | | Not at all Very  easy to collect easy to collect    1 2 3 4 5  ⬜ ⬜ ⬜ ⬜ ⬜ |
| **Time relevant components** | | |
| 16. Overall, how satisfied are you with the time that it takes to perform this test? | | Not at all Very  satisfied satisfied    1 2 3 4 5  ⬜ ⬜ ⬜ ⬜ ⬜ |
| 17. In your opinion, how many patients could be tested with this test in one 8-hour day? | | ⬜ Less than 10  ⬜ 10 to 25  ⬜ 26 to 50  ⬜ 51 to 75  ⬜ 76 to 100  ⬜ More than 100 |
| **Reading the results** | | |
| 18. Overall, how satisfied were you with the read out of the test results? | | Not at all Very  satisfied satisfied    1 2 3 4 5  ⬜ ⬜ ⬜ ⬜ ⬜ |
| 19. On a scale of 1 to 5, how difficult did you find the following: | | Not at Very  all difficult difficult  1 2 3 4 5 |
| Visibility of the control (C) band in contrast with the background | | ⬜ ⬜ ⬜ ⬜ ⬜ |
| Visibility of the test (T) band in contrast with the background | | ⬜ ⬜ ⬜ ⬜ ⬜ |
| Interpretation of the test result | | ⬜ ⬜ ⬜ ⬜ ⬜ |
| 20. Do you foresee any issues with reading the results in the lighting conditions in the settings you currently work in or have experience with? | | ⬜ Yes  ⬜ No  ⬜ Unknown  If yes, please specify: |
| 21. Has there been any color on the background of the test result or control band area that makes the interpretation of the bands difficult? | | ⬜ Yes  ⬜ No  ⬜ Unknown  If yes, please specify which color: |
| **Training and Instructions for Use** | | |
| 22. Please rate your experience reading and using the test’s Instructions for Use. | | Not at all Very easy  easy to to  understand understand  1 2 3 4 5  ⬜ ⬜ ⬜ ⬜ ⬜ |
| 23. What, if anything, would you change about the test’s Instructions for Use? | |  |
| 24. Which instructions did you refer to more: pictures or text? | | ⬜ Pictures  ⬜ Text |
| 25. Which of the following did you find most useful in learning to use this test? | | ⬜ Lecture/presentation  ⬜ Visual aids (posters, quick guides)  ⬜ Instructions for Use  ⬜ Hands on practice  ⬜ Videos  ⬜ Other, specify: _____________________ |
| **Settings of use** | | |
| 26. Do you see this test being used in its current form in your community? | | ⬜ Yes  ⬜ No  ⬜ Unknown |
| 27. [*If yes to #27]* Where do you foresee this test being used? *Select all that apply.* | | ⬜ Clinic  ⬜ Peripheral hospital/laboratory  ⬜ Reference hospital/laboratory  ⬜ At a testing site operated by staff without specific laboratory expertise  ⬜ Other, specify: _____________ |
| 28. *[If no to #27]* Which aspects should be changed to make it suitable for use in your community? | |  |
| 29. Which aspects of this test do you think might cause difficulties in day-to-day use? *Select all that apply.* | | ⬜ Hands-on time  ⬜ Total assay time to result  ⬜ Throughput  ⬜ Test results interpretation  ⬜ Overall number of steps  ⬜ Time sensitive steps  ⬜ Test design  ⬜ Quality of material  ⬜ Training requirements  ⬜ Storage conditions and stability  ⬜ Waste management requirements  ⬜ I don’t know  ⬜ None, I see no barriers for implementation  ⬜ Other, specify: __________________ |
| 30. Please give a short explanation for each of the aspects you selected above. What could be the challenges in the day-to-day use: of this test? | |  |
| 31. Do you have any final comments about this test for COVID-19? | |  |

**EASE OF USE QUESTIONNAIRE: LumiraDx SARS-CoV-2 Ag Test**

| **General questions** | |
| --- | --- |
| 1. Overall, how satisfied are you with the test? | Not at all Very  satisfied satisfied    1 2 3 4 5  ⬜ ⬜ ⬜ ⬜ ⬜ |
| 2. What do you like most about the test? |  |
| 3. What do you like least about the test? |  |
| **Kit components** | |
| 4. How satisfied are you with each of the kit components? | Not at all Very  satisfied satisfied    1 2 3 4 5 |
| External paper box | ⬜ ⬜ ⬜ ⬜ ⬜ |
| Extraction vial | ⬜ ⬜ ⬜ ⬜ ⬜ |
| Extraction vial dropper lid | ⬜ ⬜ ⬜ ⬜ ⬜ |
| Test strip | ⬜ ⬜ ⬜ ⬜ ⬜ |
| Test strip pouch | ⬜ ⬜ ⬜ ⬜ ⬜ |
| Swab for specimen collection | ⬜ ⬜ ⬜ ⬜ ⬜ |
| Instrument | ⬜ ⬜ ⬜ ⬜ ⬜ |
| 5. Which kit components do you think could be improved? *Select all that apply* | ⬜ External paper box of kit  ⬜ Extraction vial  ⬜ Extraction vial dropper lid  ⬜ Test strip  ⬜ Test strip pouch  ⬜ Swab for specimen collection  ⬜ Instrument |
| 6. How would you improve these components? |  |
| **Device design** | |
| 7. Overall, how satisfied are you with the design of the device? | Not at all Very  satisfied satisfied    1 2 3 4 5  ⬜ ⬜ ⬜ ⬜ ⬜ |
| 8. How satisfied are you with the design of the following features? | Not at all Very  satisfied satisfied    1 2 3 4 5 |
| Size of test strip | ⬜ ⬜ ⬜ ⬜ ⬜ |
| Size of the test strip application area | ⬜ ⬜ ⬜ ⬜ ⬜ |
| Size of the instrument | ⬜ ⬜ ⬜ ⬜ ⬜ |
| LumiraDx instrument interface/result display | ⬜ ⬜ ⬜ ⬜ ⬜ |
| Logical sequence of steps | ⬜ ⬜ ⬜ ⬜ ⬜ |
| 9. Which design features do you think could be improved? *Select all that apply* | ⬜ Size of test strip  ⬜ Size of the test strip sample application area  ⬜ Size of the reader  ⬜ LumiraDx reader interface/result display  ⬜ Logical sequence of steps  ⬜ Other, specify: ______________ |
| 10. How would you improve these components? |  |
| **Ease of use** | |
| 11. On a scale of 1 to 5, please rate the ease of using this test. | Not at all Very  easy to use easy to use    1 2 3 4 5  ⬜ ⬜ ⬜ ⬜ ⬜ |
| 12. On a scale of 1 to 5, how difficult were the following steps? | Not at Very  all difficult difficult  1 2 3 4 5 |
| Instrument set-up | ⬜ ⬜ ⬜ ⬜ ⬜ |
| Check expiry date | ⬜ ⬜ ⬜ ⬜ ⬜ |
| Open the seal of the extraction vial | ⬜ ⬜ ⬜ ⬜ ⬜ |
| Insert the swab into the tube | ⬜ ⬜ ⬜ ⬜ ⬜ |
| Ease of swab extraction procedure | ⬜ ⬜ ⬜ ⬜ ⬜ |
| Ability to perform extraction procedure consistently | ⬜ ⬜ ⬜ ⬜ ⬜ |
| Remove the test strip from the pouch | ⬜ ⬜ ⬜ ⬜ ⬜ |
| Insert the test strip into the instrument |  |
| Apply the exact quantity of the sample onto the test strip | ⬜ ⬜ ⬜ ⬜ ⬜ |
| 13. Have you had any challenges with using this test? | ⬜ Yes  ⬜ No  If yes, please explain: |
| **Time relevant components** | |
| 14. Overall, how satisfied are you with the time that it takes to perform this test? | Not at all Very  satisfied satisfied    1 2 3 4 5  ⬜ ⬜ ⬜ ⬜ ⬜ |
| 15. In your opinion, how many patients could be tested with this test in one 8-hour day? | ⬜ Less than 10  ⬜ 10 to 25  ⬜ 26 to 50  ⬜ 51 to 75  ⬜ 76 to 100  ⬜ More than 100 |
| **Reading the results** | |
| 16. Overall, how satisfied were you with the read out of the test results? | Not at all Very  satisfied satisfied    1 2 3 4 5  ⬜ ⬜ ⬜ ⬜ ⬜ |
| 17. Do you foresee any issues with reading the results in the lighting conditions in the settings you currently work in or have experience with? | ⬜ Yes  ⬜ No  ⬜ Unknown  If yes, please specify: |
| **Training and Instructions for Use** | |
| 18. Please rate your experience reading and using the test’s Instructions for Use. | Not at all Very easy  easy to to  understand understand  1 2 3 4 5  ⬜ ⬜ ⬜ ⬜ ⬜ |
| 19. What, if anything, would you change about the test’s Instructions for Use? |  |
| 20. Which instructions did you refer to more: pictures or text? | ⬜ Pictures  ⬜ Text |
| 21. Which of the following did you find most useful in learning to use this test? | ⬜ Lecture/presentation  ⬜ Visual aids (posters, quick guides)  ⬜ Instructions for Use  ⬜ Hands on practice  ⬜ Videos  ⬜ Other, specify: _____________________ |
| **Settings of use** | |
| 22. Do you see this test being used in its current form in your community? | ⬜ Yes  ⬜ No  ⬜ Unknown |
| 23. [*If yes to #23]* Where do you foresee this test being used? | ⬜ Clinic  ⬜ Peripheral hospital/laboratory  ⬜ Reference hospital/laboratory  ⬜ At a testing site operated by staff without specific laboratory expertise  ⬜ Other, specify: _____________ |
| 24. *[If no to #23]* Which aspects should be changed to make it suitable for use in your community? |  |
| 25. Which aspects of this test do you think might cause difficulties in day-to-day use? *Select all that apply.* | ⬜ Hands-on time  ⬜ Battery life of instrument  ⬜ Total assay time to result  ⬜ Batch processing  ⬜ Throughput  ⬜ Test results interpretation  ⬜ Overall number of steps  ⬜ Time sensitive steps  ⬜ Test design  ⬜ Quality of material  ⬜ Training requirements  ⬜ Storage conditions and stability  ⬜ Waste management requirements  ⬜ I don’t know  ⬜ None, I see no barriers for implementation  ⬜ Other, specify: __________________ |
| 26. Please give a short explanation for each of the aspects you selected above. What could be the challenges in the day-to-day use: of this test? |  |
| 27. Do you have any final comments about this test for COVID-19? |  |

**Supplementary Material E.** Matrix for Ease of Use Assessment Analysis.

| **#** | **Question** | **Satisfactory** | **Satisfactory** | **Average** | **Average** | **Unsatisfactory** | **Unsatisfactory** |
| --- | --- | --- | --- | --- | --- | --- | --- |
| 1 | Participant ID | - | - | - | - | - | - |
| 2 | Date | - | - | - | - | - | - |
| 3 | Occupation | - | - | - | - | - | - |
| 4 | Level of education | - | - | - | - | - | - |
| 5 | Years of relevant experience | - | - | - | - | - | - |
| 6 | Which test are you assessing? | - | - | - | - | - | - |
| 7 | How many times have you performed this test? | - | - | - | - | - | - |
| 8 | Overall, how satisfied are you with the test? | very satisfied (5) | 4 | 3 | - | 2 | not at all satisfied (1) |
| 9 | What do you like most about the test? | - | - | - | - | - | - |
| 10 | What do you like least about the test? | - | - | - | - | - | - |
| 11 | How satisfied are you with each of the kit components? [external paper box] | very satisfied (5) | 4 | 3 | - | 2 | not at all satisfied (1) |
| 12 | How satisfied are you with each of the kit components? [extraction buffer tube/ vial] | very satisfied (5) | 4 | 3 | - | 2 | not at all satisfied (1) |
| 13 | How satisfied are you with each of the kit components? [extraction buffer nozzle cap/ extraction vial dropper lid] | very satisfied (5) | 4 | 3 | - | 2 | not at all satisfied (1) |
| 14 | How satisfied are you with each of the kit components? [buffer tube rack] | very satisfied (5) | 4 | 3 | - | 2 | not at all satisfied (1) |
| 15 | How satisfied are you with each of the kit components? [swab] | very satisfied (5) | 4 | 3 | - | 2 | not at all satisfied (1) |
| 16 | How satisfied are you with each of the kit components? [test device/ strip] | very satisfied (5) | 4 | 3 | - | 2 | not at all satisfied (1) |
| 17 | How satisfied are you with each of the kit components? [test device/strip foil pouch] | very satisfied (5) | 4 | 3 | - | 2 | not at all satisfied (1) |
| 18 | How satisfied are you with each of the kit components? [positive/negative control swab] | very satisfied (5) | 4 | 3 | - | 2 | not at all satisfied (1) |
| 19 | How satisfied are you with each of the kit components? [Instrument] | very satisfied (5) | 4 | 3 | - | 2 | not at all satisfied (1) |
| 20 | Which kit components do you think could be improved? Select all that apply. | - | - | - | - | - | - |
| 21 | For each selected, how would you improve this component? | - | - | - | - | - | - |
| 22 | Overall, how satisfied are you with the design of the device? | very satisfied (5) | 4 | 3 | - | 2 | not at all satisfied (1) |
| 23 | How satisfied are you with the design of the following features? [size of the test device/strip] | very satisfied (5) | 4 | 3 | - | 2 | not at all satisfied (1) |
| 24 | How satisfied are you with the design of the following features? [size of the well/application area to add the extracted sample] | very satisfied (5) | 4 | 3 | - | 2 | not at all satisfied (1) |
| 25 | How satisfied are you with the design of the following features? [size of the reading window] | very satisfied (5) | 4 | 3 | - | 2 | not at all satisfied (1) |
| 26 | How satisfied are you with the design of the following features? [size of the instrument] | very satisfied (5) | 4 | 3 | - | 2 | not at all satisfied (1) |
| 27 | How satisfied are you with the design of the following features? [instrument interface/result display] | very satisfied (5) | 4 | 3 | - | 2 | not at all satisfied (1) |
| 28 | How satisfied are you with the design of the following features? [logical sequence of steps] | very satisfied (5) | 4 | 3 | - | 2 | not at all satisfied (1) |
| 29 | Which design features do you think could be improved? Select all that apply. | - | - | - | - | - | - |
| 30 | For each selected, how would you improve this feature? | - | - | - | - | - | - |
| 31 | Please rate the ease of using this test. | very easy to use (5) | 4 | 3 | - | 2 | not at all easy to use (5) |
| 32 | How difficult were the following steps? [instrument set up] | not at all difficult (1) | 2 | 3 | - | 4 | very difficult (5) |
| 33 | How difficult were the following steps? [check expiry date] | not at all difficult (1) | 2 | 3 | - | 4 | very difficult (5) |
| 34 | How difficult were the following steps? [remove the test device/ test strip from the pouch] | not at all difficult (1) | 2 | 3 | - | 4 | very difficult (5) |
| 35 | How difficult were the following steps? [open the extraction buffer tube/ open seal of extraction vial] | not at all difficult (1) | 2 | 3 | - | 4 | very difficult (5) |
| 36 | How difficult were the following steps? [insert the swab into the tube] | not at all difficult (1) | 2 | 3 | - | 4 | very difficult (5) |
| 37 | How difficult were the following steps? [ease of swab extraction procedure] | not at all difficult (1) | 2 | 3 | - | 4 | very difficult (5) |
| 38 | How difficult were the following steps? [ability to perform extraction procedure consistently] | not at all difficult (1) | 2 | 3 | - | 4 | very difficult (5) |
| 39 | How difficult were the following steps? [insert the test strip into the instrument] | not at all difficult (1) | 2 | 3 | - | 4 | very difficult (5) |
| 40 | How difficult were the following steps? [transfer the correct number of drops into the sample well of the test device/ apply the exact quantity of the sample onto the test strip] | not at all difficult (1) | 2 | 3 | - | 4 | very difficult (5) |
| 41 | Have you had any challenges with using this test? | - | - | - | - | - | - |
| 42 | If yes, please explain. | - | - | - | - | - | - |
| 43 | Please rate your experience collecting participant samples for this test, if applicable. | very easy to collect (5) | 2 | 3 | - | 4 | not at all easy to collect (5) |
| 44 | Overall, how satisfied are you with the time it takes to perform this test? | very satisfied (5) | 4 | 3 | - | 2 | not at all satisfied (1) |
| 45 | In your opinion, how many patients could be tested with this test in one 8-hour day? | >100 | - | 51 to 100 | 10 to 50 | - | <10 |
| 46 | Overall, how satisfied were you with the read out of the test results? | very satisfied (5) | 4 | 3 | - | 2 | not at all satisfied (1) |
| 47 | How difficult did you find the following? [visibility of the control band in contrast with the background] | not at all difficult (1) | 2 | 3 | - | 4 | very difficult (5) |
| 48 | How difficult did you find the following? [visibility of the test band in contrast with the background] | not at all difficult (1) | 2 | 3 | - | 4 | very difficult (5) |
| 49 | How difficult did you find the following? [interpretation of the test result] | not at all difficult (1) | 2 | 3 | - | 4 | very difficult (5) |
| 50 | Do you foresee any issues with reading the results in the lighting conditions in the settings you currently work in or have experience with? | No | - | - | - | - | Yes |
| 51 | If yes, please explain. | - | - | - | - | - | - |
| 52 | Has there been any color on the background of the test result or control band area that makes the interpretation of the bands difficult? | - | - | - | - | - | - |
| 53 | If yes, please specify which color. | - | - | - | - | - | - |
| 54 | Please rate your experience reading and using the test's Instructions for Use. | very easy to understand (5) | 4 | 3 | - | 2 | not at all easy to understand (1) |
| 55 | What, if anything, would you change about the test's Instructions for Use? | - | - | - | - | - | - |
| 56 | Which instructions did you refer to more: pictures or text? | - | - | - | - | - | - |
| 57 | Which of the following did you find most useful in learning to use the test (select all that apply). | - | - | - | - | - | - |
| 58 | Do you see this test being used in its current form in your community? | Yes | - | - | - | - | No |
| 59 | Where do you foresee this test being used? Select all that apply. | At a testing site operated by staff without laboratory expertise | Clinic | Peripheral hospital | - | Reference hospital | None |
| 60 | Which aspects should be changed to make it suitable for use in your community? | 0 | - | 1 to 2 | - | >2 | - |
| 61 | Which aspects of this test do you think might cause difficulty in day-to-day use? Select all that apply. | 0 | - | 1 | 2 | 3 | 4 or more |
| 62 | Please give a short explanation for each aspect listed above. What could be the challenges in day-to-day use of the test? | - | - | - | - | - | - |
| 63 | Do you have any final comments about this test for COVID-19? | - | - | - | - | - | - |

**Supplementary Material F**. Vaccination status of study participants, by group.

|  | **Index cases** | **Close contacts,**  **non-household** | | **Close contacts,**  **household** | |
| --- | --- | --- | --- | --- | --- |
| **PCR results*** | Pos | Neg | Pos | Neg | Pos |
| **Total N** | 50 | 122 | 28 | 28 | 37 |
| **Fully vaccinated**** | **N=12** | **N=34** | **N=9** | **N=7** | **N=8** |
| **Vaccine type** |  |  |  |  |  |
| AstraZeneca | 2 | 7 | 3 | 1 | 0 |
| CoronaVac | 8 | 21 | 4 | 4 | 5 |
| Johnson & Johnson | 2 | 2 | 2 | 0 | 0 |
| Pfizer | 0 | 4 | 0 | 2 | 3 |
| **Partially vaccinated** | **N=24** | **N=59** | **N=10** | **N=14** | **N=11** |
| **Vaccine type** |  |  |  |  |  |
| AstraZeneca | 17 | 32 | 5 | 7 | 3 |
| CoronaVac | 1 | 1 | 2 | 0 | 1 |
| Johnson & Johnson | 0 | 0 | 0 | 0 | 0 |
| Pfizer | 6 | 26 | 3 | 7 | 7 |
| **Unvaccinated** | **N=14** | **N=29** | **N=9** | **N=6** | **N=18** |

* For household contacts, PCR positive indicates that a participant was PCR positive at any visit.

** Fully vaccinated classification indicates that a participant had received all required vaccine doses and was >14 days since receipt of the last vaccine dose at enrollment.

**Supplementary Material G**. Viral load value relationships among close contacts by vaccination status and infection category.

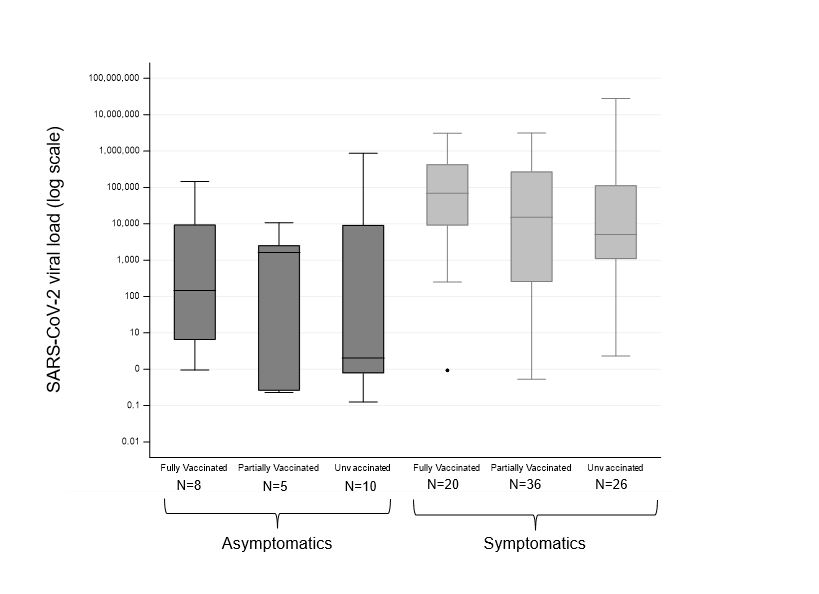

**Supplementary Material H.** SARS-CoV-2 strains detected in sequenced nasopharyngeal specimens, over time.

SARS-CoV-2 strains detected in sequenced nasopharyngeal specimens over time

\# of specimens (cumulative)

| Date of sample collection | Gamma, V3 | Delta |
| --- | --- | --- |
| 2021-07-06 | 1 | 0 |
| 2021-07-07 | 2 | 0 |
| 2021-07-08 | 6 | 0 |
| 2021-07-12 | 7 | 0 |
| 2021-07-13 | 10 | 0 |
| 2021-07-14 | 12 | 0 |
| 2021-07-15 | 13 | 0 |
| 2021-07-16 | 15 | 0 |
| 2021-07-17 | 16 | 0 |
| 2021-07-18 | 17 | 0 |
| 2021-07-19 | 18 | 0 |
| 2021-07-20 | 19 | 0 |
| 2021-07-21 | 20 | 0 |
| 2021-07-22 | 21 | 0 |
| 2021-07-23 | 26 | 0 |
| 2021-07-26 | 28 | 0 |
| 2021-07-27 | 31 | 0 |
| 2021-07-28 | 37 | 0 |
| 2021-07-29 | 39 | 0 |
| 2021-08-03 | 39 | 3 |
| 2021-08-04 | 41 | 3 |
| 2021-08-05 | 45 | 3 |
| 2021-08-06 | 46 | 3 |
| 2021-08-09 | 47 | 3 |
| 2021-08-10 | 48 | 3 |
| 2021-08-11 | 50 | 3 |
| 2021-08-12 | 53 | 3 |
| 2021-08-18 | 57 | 3 |
| 2021-08-20 | 58 | 4 |
| 2021-08-22 | 59 | 4 |
| 2021-08-24 | 62 | 4 |
| 2021-08-25 | 65 | 4 |
| 2021-08-26 | 66 | 4 |
| 2021-09-02 | 67 | 4 |
| 2021-09-08 | 67 | 5 |
| 2021-09-10 | 68 | 5 |
| 2021-09-11 | 68 | 6 |
| 2021-09-15 | 68 | 8 |
| 2021-09-16 | 68 | 10 |
| 2021-09-21 | 68 | 11 |
| 2021-09-24 | 68 | 12 |
| 2021-09-27 | 68 | 15 |
| 2021-09-29 | 68 | 16 |

**Supplementary Material I.** Descriptive plots for all close contacts positive by the RT-PCR reference assay with multiple visits**.** Visit numbers are presented on the x-axis, and test results and symptom status are shown on the y-axis. Symptom status is presented independently of the RT-PCR reference assay result.
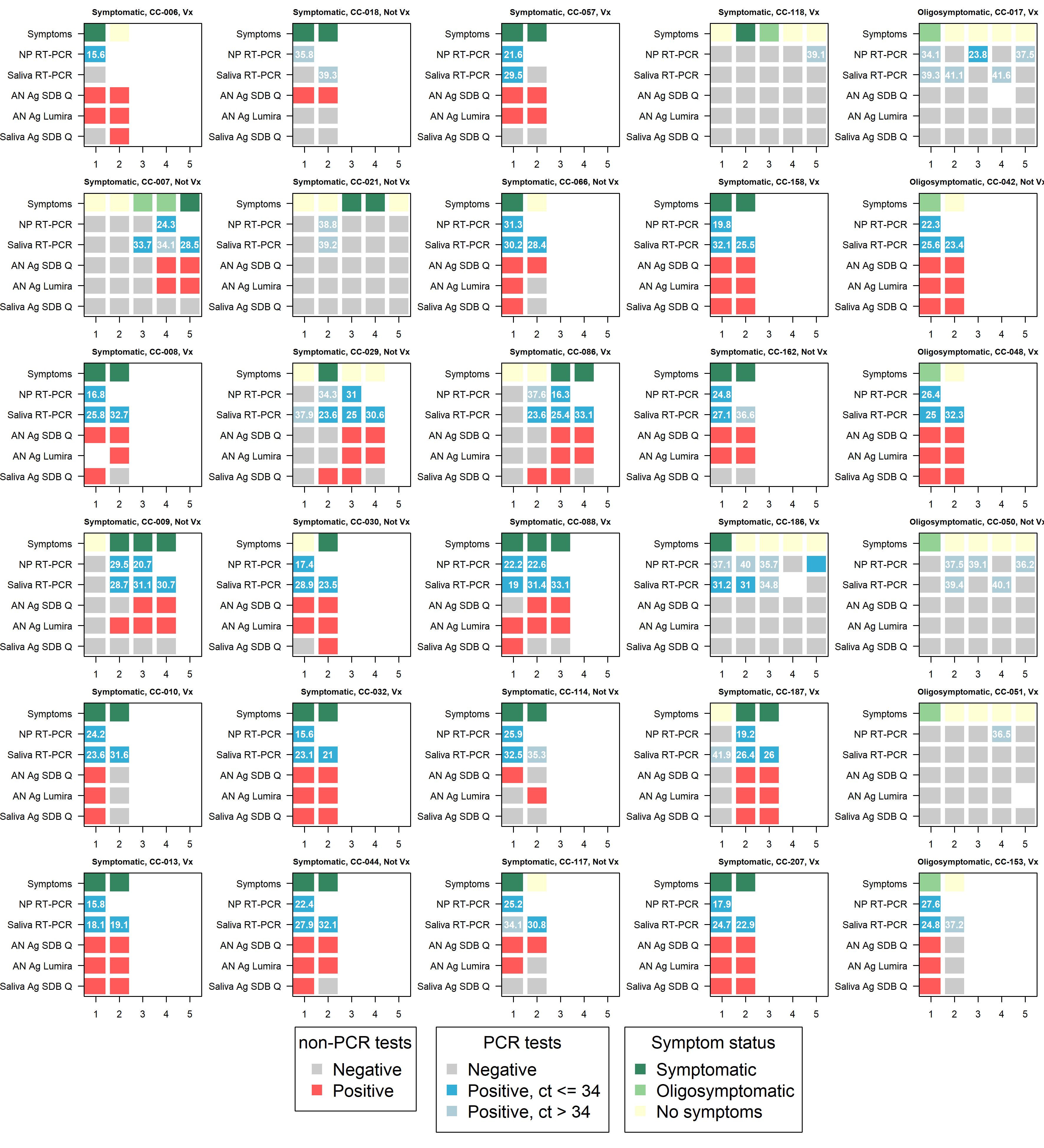

**Supplementary Material J.** Usability study analysis.

a. Participant demographics

|  | **STANDARD Q Nasal**  **N (%)** | **STANDARD Q Saliva**  **N (%)** | **LumiraDx**  **N (%)** | **TOTAL**  **N (%)** |
| --- | --- | --- | --- | --- |
| **Total participants** | 11 (100%) | 3 (100%) | 5 (100%) | 12 (100%) |
| **Occupation** |  |  |  |  |
| Laboratory technician | 2 (18%) | 3 (100%) | 3 (60%) | 3 (25%) |
| Physician | 0 (0%) | 0 (0%) | 0 (0%) | 0 (0%) |
| Registered nurse | 1 (9%) | 0 (0%) | 0 (0%) | 1 (8%) |
| Field agent | 8 (73%) | 0 (0%) | 2 (40%) | 8 (67%) |
| Other | 0 (0%) | 0 (0%) | 0 (0%) | 0 (0%) |
| **Highest level of education** |  |  |  |  |
| High school | 1 (9%) | 0 (0%) | 0 (0%) | 1 (8%) |
| Technical certificate | 0 (0%) | 0 (0%) | 0 (0%) | 0 (0%) |
| University, bachelor | 8 (73%) | 2 (67%) | 3 (60%) | 9 (75%) |
| University, master | 0 (0%) | 0 (0%) | 0 (0%) | 0 (0%) |
| University, doctorate | 0 (0%) | 0 (0%) | 0 (0%) | 0 (0%) |
| Physician | 0 (0%) | 0 (0%) | 0 (0%) | 0 (0%) |
| Other (higher education) | 1 (9%) | 0 (0%) | 1 (20%) | 1 (8%) |
| Prefer not to respond | 1 (9%) | 1 (33%) | 1 (20%) | 1 (8%) |
| **Years of relevant experience** |  |  |  |  |
| Less than 1 year | 9 (82%) | 0 (0%) | 2 (40%) | 9 (75%) |
| 1 to 3 years | 0 (0%) | 0 (0%) | 0 (0%) | 0 (0%) |
| 3 to 5 years | 1 (9%) | 2 (67%) | 2 (40%) | 2 (17%) |
| 5 to 10 years | 1 (9%) | 1 (33%) | 1 (20%) | 1 (8%) |
| More than 10 years | 0 (0%) | 0 (0%) | 0 (0%) | 0 (0%) |
| Prefer not to respond | 0 (0%) | 0 (0%) | 0 (0%) | 0 (0%) |
| **How many times performed** |  |  |  |  |
| 1 to 10 | 2 (18%) | 0 (0%) | 2 (40%) | - |
| 11 to 50 | 4 (36%) | 1 (33%) | 0 (0%) | - |
| 51 to 100 | 1 (9%) | 0 (0%) | 2 (40%) | - |
| >100 | 4 (36%) | 2 (67%) | 1 (20%) | - |

b. STANDARD Q COVID-19 Ag Test: Nasal

| **Interpretation key** | |
| --- | --- |
|  | Satisfactory |
|  | Average |
|  | Unsatisfactory |

| **STANDARD Q COVID-19 Ag Test: Nasal (SD Biosensor, Republic of Korea)** | | | | | | | | | |
| --- | --- | --- | --- | --- | --- | --- | --- | --- | --- |
| **General** | Overall satisfaction with test |  | On a system usability scale, the test scored **79.8** out of 100 points. | | | | | | |
|  | Ease of reading/using IFU |  | Challenges mainly related to dropping the correct number of drops onto the application area; several respondents also noted a preference for a larger swab and a sturdier buffer tube rack. Users reported that the time from specimen collection to results and storage conditions in hot/humid settings may cause barriers to use. | | | | | | |
| **Quality of the test's hardware** | External paper box |  |  | **Ease of test execution** | Overall ease of use of test |  |  | **Procedure time** | Overall satisfaction with time requirements |
|  | Extraction buffer tube/vial |  |  |  | Check expiry date |  |  |  | 8-hour throughput |
|  | Extraction buffer nozzle cap/extraction vial dropper lid |  |  |  | Remove the test device/test strip from the pouch |  |  | **Ease of result interpretation** | Overall satisfaction with read-out |
|  | Buffer tube rack |  |  |  | Open the extraction buffer tube/open seal of extraction vial |  |  |  | Issues with lighting |
|  | Swab for specimen collection |  |  |  | Insert the swab into the tube |  |  |  | Visibility of the control (C) band |
|  | Test device/strip |  |  |  | Swab extraction procedure |  |  |  | Visibility of the test (T) band |
|  | Test device/strip foil pouch |  |  |  | Perform extraction procedure consistently |  |  |  | Interpretation of the test results |
|  | Positive/negative control swab |  |  |  | Transfer the correct number of drops into the sample well of the test device/apply the exact quantity of the sample onto the test strip |  |  | **Fields of application** | Appropriateness for use in community |
|  | Size of the test device/strip |  |  |  | Ease of sample collection (if applicable) |  |  |  | Batch testing possible |
|  | Size of the well/application area to add the extracted sample |  |  |  | Logical sequence of steps |  |  |  | Settings of use |
|  | Size of the reading window |  |  |  |  |  |  |  | Recommended changes for use |
|  | Overall design |  |  |  |  |  |  |  | Aspects causing difficulty for use |

c. STANDARD Q COVID-19 Ag Test: Saliva

| **STANDARD Q COVID-19 Ag Test: Saliva (SD Biosensor, Republic of Korea)** | | | | | | | | |  |
| --- | --- | --- | --- | --- | --- | --- | --- | --- | --- |
| **General** | Overall satisfaction with test |  | On a system usability scale, the test scored **77.5** out of 100 points. | | | | | | |
|  | Ease of reading/using IFU |  | Respondents main concern was the test’s performance. One user also expressed that interpreting weak positive results may be challenging in low light. Users reported that the storage conditions and stability in hot and humid settings may cause barriers to use. | | | | | | |
| **Quality of the test's hardware** | External paper box |  |  | **Ease of test execution** | Overall ease of use of test |  |  | **Procedure time** | Overall satisfaction with time requirements |
|  | Extraction buffer tube/vial |  |  |  | Check expiry date |  |  |  | 8-hour throughput |
|  | Extraction buffer nozzle cap/ extraction vial dropper lid |  |  |  | Remove the test device/test strip from the pouch |  |  | **Ease of result interpretation** | Overall satisfaction with read-out |
|  | Buffer tube rack |  |  |  | Open the extraction buffer tube/open seal of extraction vial |  |  |  | Issues with lighting |
|  | Swab for specimen collection |  |  |  | Insert the swab into the tube |  |  |  | Visibility of the control (C) band |
|  | Test device/strip |  |  |  | Swab extraction procedure |  |  |  | Visibility of the test (T) band |
|  | Test device/strip foil pouch |  |  |  | Perform extraction procedure consistently |  |  |  | Interpretation of the test results |
|  | Positive/negative control swab |  |  |  | Transfer the correct number of drops into the sample well of the test device/ apply the exact quantity of the sample onto the test strip |  |  | **Fields of application** | Appropriateness for use in community |
|  | Size of the well/application area to add the extracted sample |  |  |  | Logical sequence of steps |  |  |  | Batch testing possible |
|  | Size of the reading window |  |  |  |  |  |  |  | Settings of use |
|  | Overall design |  |  |  |  |  |  |  | Recommended changes for use |
|  |  |  |  |  |  |  |  |  | Aspects causing difficulty for use |

d. LumiraDx SARS-CoV-2 Ag Test

| **SARS-CoV-2 Ag Test (LumiraDx™ Limited, United Kingdom)** | | | | | | | | | |
| --- | --- | --- | --- | --- | --- | --- | --- | --- | --- |
| **General** | Overall satisfaction with test |  | On a system usability scale, the test scored **83.5** out of 100 points. | | | | | | |
|  | Ease of reading/using IFU |  | Reported challenges included operating errors, processing time, and the number of test strips included per kit. 3/5 users suggested increasing the size of the sample application area. Users reported that the hands-on time (1/5), the battery life of the instrument (2/5), the total assay time to result (1/5), inability to batch process (1/5), number of time sensitive steps (1/5), training requirements (1/5), and storage conditions/stability requirements (1/5) may cause challenges/ barriers to use. | | | | | | |
| **Quality of the test's hardware** | External paper box |  |  | **Ease of test execution** | Remove the test device/test strip from the pouch |  |  | **Procedure time** | Overall satisfaction with time requirements |
|  | Extraction buffer tube/vial |  |  |  | Open the extraction buffer tube/open seal of extraction vial |  |  |  | 8-hour throughput |
|  | Extraction buffer nozzle cap/extraction vial dropper lid |  |  |  | Insert the swab into the tube |  |  | **Ease of result interpretation** | Overall satisfaction with read-out |
|  | Swab for specimen collection |  |  |  | Swab extraction procedure |  |  |  | Issues with lighting |
|  | Test device/strip |  |  |  | Perform extraction procedure consistently |  |  | **Fields of application** | Appropriateness for use in community |
|  | Test device/strip foil pouch |  |  |  | Insert the test strip into the instrument |  |  |  | Batch testing possible |
|  | Instrument design |  |  |  | Transfer the correct number of drops into the sample well of the test device/apply the exact quantity of the sample onto the test strip |  |  |  | Settings of use |
|  | Size of the test device/strip |  |  |  | Logical sequence of steps |  |  |  | Recommended changes for use |
|  | Size of the well/application area to add the extracted sample |  |  |  |  |  |  |  | Aspects causing difficulty for use |
|  | Size of the instrument |  |  |  |  |  |  |  |  |
|  | Instrument interface/result display |  |  |  |  |  |  |  |  |
|  | Overall design |  |  |  |  |  |  |  |  |
